# A systematic review exploring the association between the human gut microbiota and brain connectivity in health and disease

**DOI:** 10.1101/2022.11.29.22282879

**Authors:** Danique Mulder, Esther Aarts, Alejandro Arias Vasquez, Mirjam Bloemendaal

## Abstract

A body of pre-clinical evidence shows how the gut microbiota influence brain functioning, including brain connectivity. A growing number of studies have investigated the association between the gut microbiota and brain connectivity in humans. Linking brain connectivity measures to the gut microbiota can provide important mechanistic insights into the bi-directional gut-brain communication. In this systematic review, we therefore synthesized the available literature assessing this association, evaluating the degree of consistency in microbiota-connectivity associations. Following the PRISMA guidelines, a PubMed search was conducted, including studies published up to September 1, 2022. We identified 16 studies that met the inclusion criteria. Several bacterial genera, including *Prevotella*, *Bacteroides*, *Ruminococcus* and *Blautia* were most consistently reported in association with brain connectivity. Additionally, the salience (specifically the insula and anterior cingulate cortex), default mode and frontoparietal networks were most frequently associated with the gut microbiota, both in terms of microbial diversity and composition. Altogether, based on our synthesis, there is evidence for an association between the gut microbiota and brain connectivity. However, the specificity of the signal is yet unclear as most findings were poorly replicated across studies. The current studies show substantial inter-study heterogeneity in methodology and reporting, limiting the robustness and reproducibility of the findings and emphasizing the need to harmonize methodological approaches. To enhance comparability and replicability, future research should focus on further standardizing processing pipelines and employ data-driven multivariate analysis strategy.

## 2 Introduction

Neuronal connections are determined by the distance between neurons. Spatially closer neurons have a higher probability of being connected than those further away. As a result, the structural wiring of the brain represents a complex network with clusters of highly connected regions (i.e., structural connectivity) [1] that provide a basis for functional communication between brain areas (i.e., functional connectivity) as measured by temporal coincidence of neuronal activation patterns of anatomically separated brain regions [2]. Patterns of communication between fixed sets of brain regions form functional networks [3, 4], which can be identified both during active cognitive tasks and during rest.

Functional networks measured during rest (resting-state networks) reflect, among others, processes related to cognitive functions. For example, the salience network (SN) is involved in the detection of behaviorally relevant stimuli [5], the frontoparietal network (FPN) is involved in the coordination of cognitive control [6], and the default mode network (DMN) is linked to basal, stimulus-independent cognitive processes such as information integration and mind-wandering [7]. In addition to cognitive functioning, connectivity may also reflect intrinsic processes, such as emotional and interoceptive awareness [8, 9]. Dysfunction in connectivity networks is observed in a range of psychiatric and neurodevelopmental disorders, including Attention Deficit/Hyperactivity Disorder [10] and Schizophrenia [11], in certain cases even prior to diagnosis [12].

The functional and structural connectivity patterns of our brain are affected by numerous genetic and non-genetic interacting factors. Our genetic makeup has a significant biological effect on both brain structure and function: heritability studies have shown that the additive genetic contribution explains approximately 50% to 93% of the variance in structural connectivity [13] and 20% to 40% of the variance in functional connectivity [14]. Likewise, the brain can change under the influence of environmental factors. Multiple studies have reported changes in the connectivity strength following a mindfulness training, both in structural [15] and in functional connectivity [16]. Additionally, a systematic review concluded that diet (specifically a lower quality diet) was related to changes in both structural and functional connectivity throughout the brain [17].

Such environmental factors, including diet, may exert their influence on the brain through the gut-brain axis (GBA), among others via modulation of the gut microbiota [18]. The GBA refers to the bidirectional communication super highway connecting the gastro-intestinal system with the central nervous system (CNS) through endocrine, immune, and neural/vagal pathways [19]. The gut microbiota, comprising the trillions of microbes (predominantly bacteria) residing in the intestines, can modulate gut-brain communication, for example through the production of neuroactive metabolites, and by maintaining the gastro-intestinal and blood-brain barriers [19].

A majority of the studies investigating the (microbiota-)gut-brain axis (MGBA) in humans focus on behavioral measures, including clinical diagnoses and questionnaires, providing evidence for a link between the gut microbiota composition and cognitive and emotional functioning [20–22]. In recent years the number of studies incorporating neuroimaging into the microbiota-gut-brain investigation has also increased rapidly. Specifically, the acquisition of functional and structural connectivity data is relatively standardized and often done in rest (i.e., without task instructions), making it a suitable research method for many participant populations, including children and patients. Linking such connectivity measures to the gut microbiota can provide important mechanistic insights into the bi-directional gut-brain communication. Therefore, we herein systematically review the available studies associating the gut microbiota with brain connectivity, in an effort to evaluate the degree of consistency in this association.

## 3 Method

### 3.1 Search strategy

A systematic search on the PubMed database was conducted for reports published up to September 1, 2022. The aim was to capture all human studies that 1) collected a fecal sample to assess the gut microbiota, 2) assessed *in vivo* functional or structural brain connectivity, and 3) performed statistical analysis on the association between the gut microbiota and brain connectivity. Only peer-reviewed original research studies published in English were included. Two independent raters (DM, MB) reviewed the titles and abstracts and came to a consensus about study inclusion. After inclusion, the following data was independently extracted by two authors (DM, MB): demographics, sample characteristics, method of gut microbiota estimation, method of brain connectivity estimation, statistical methods and relevant results. Details on the search strategy and study inclusion are provided in **Fig. 1.**

**Figure 1.**
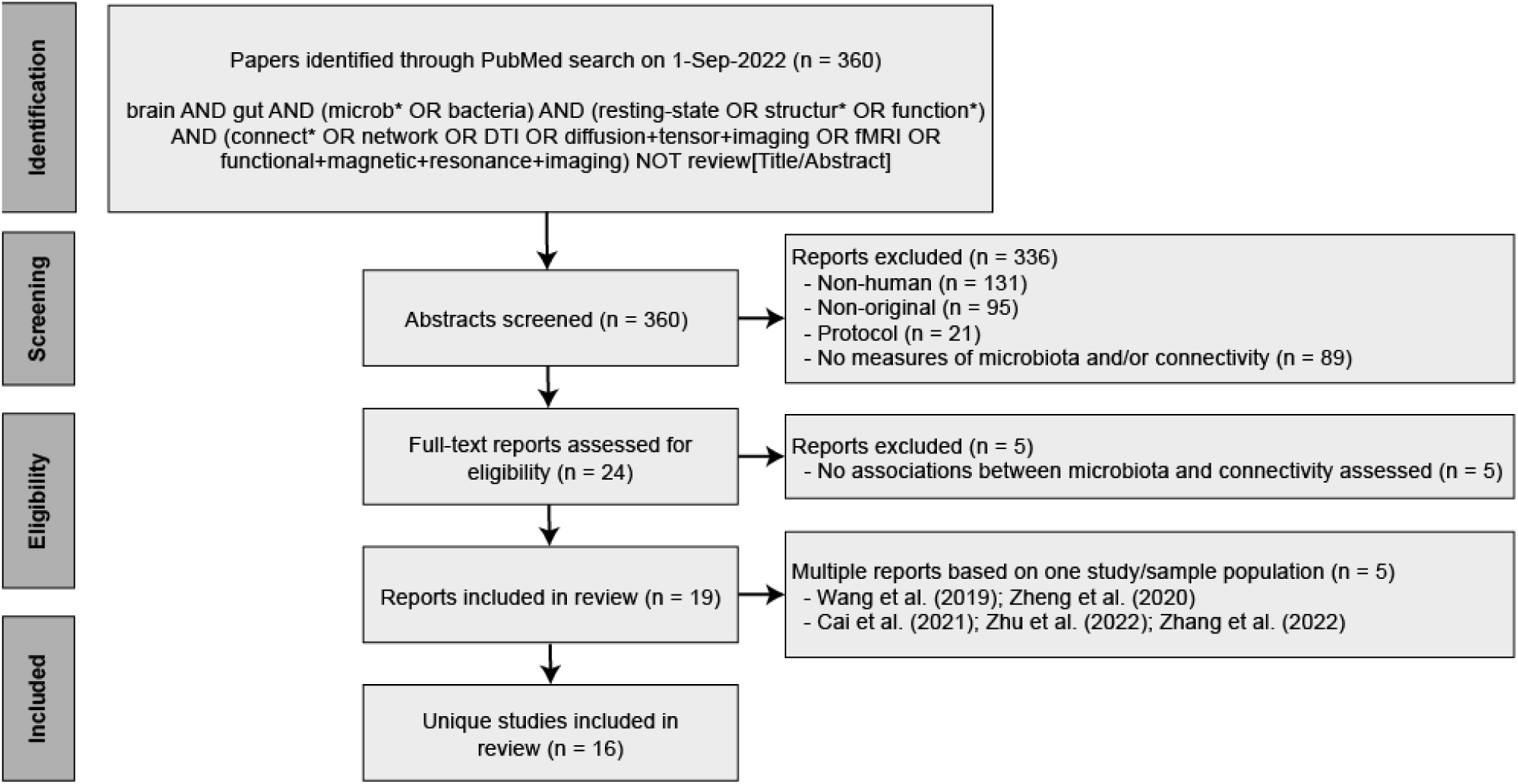
PRISMA flow diagram detailing the database search, number of reports screened, and number of studies included.

### 3.2 Quality assessment

Two authors (DM, MB) assessed the risk of bias in the included studies using the National Institutes of Health (NIH) National Health, Lung and Blood Institute Study Quality Assessment Tool for observational Cohort and Cross-sectional Studies [23]. The tool was modified to also be suitable for case-control and before-after studies with no control group (specified in **Table S3**). We used the method section of the STORMS checklist (v1.03) [24] to assess gut microbiota measurements, and an adaptation of the guidelines published by Poldrack et al. [25] to assess brain connectivity measurements (**Table S4-5**). Study quality was rated as ‘Good’ for assessments of 75% or higher; ‘Fair’ for assessments between 50% and 75%; or ‘Poor’ for assessments lower than 50%.

## 4 Results

A comprehensive literature search on PubMed yielded 360 reports, of which a total of 19 publications based on 16 unique studies met the inclusion criteria (**Fig. 1**). Two of the included studies produced more than one publication ([26–28] and [29, 30]). For those studies, the findings reported in the individual publications were pooled.

### 4.1 Population characteristics (***table 1***)

There was a wide variation in the target populations. Eight studies were conducted in healthy individuals [26–28,31–36] including a study conducted in smokers [31], in newborns [35] and in infants [34]. The other eight studies were performed in a range of disease populations, including: bipolar disorder [37], cirrhosis [38], end stage renal disease [29, 30], irritable bowel syndrome [39, 40], major depressive disorder [41], obesity [42] and patients undergoing a laparoscopic/vertical sleeve gastrectomy [43, 44]. Four of those studies were case-controlled [29,30,37,40,42] and three were longitudinal [39,43,44]. All other studies performed associations between gut microbiota composition and brain connectivity based on a single group and timepoint. Further characteristics of the study populations are listed in **Table 1**.

**Table 1.** Overview of population characteristics

|  | Ref | First author (year) | Country | Target population | Sample size | % female | Age in years (SD) | Antibiotic use | Pre-/ pro-/ synbiotic use | Quality Rating | Other |
| --- | --- | --- | --- | --- | --- | --- | --- | --- | --- | --- | --- |
| **Healthy population Adults** | 31 | Curtis (2019) | USA | Healthy (smokers / eCig users / non-smokers) | 30* | 7% | 33 (2.7) |  |  | Poor | Smoking groups pooled in analyses |
|  | 26, 27, 28 | Cai (2021) / Zhu (2022) / Zhang (2022) | China | Healthy | 157 | 49% | 22.3 (2.4) | No antibiotics month before inclusion |  | Fair |  |
|  | 32 | Hall (2021) | Australia | Healthy | 38% | 60.5% | 31.7 (9.7) | No antibiotics 3 months before inclusion | No probiotics 3 months before inclusion | Fair |  |
|  | 33 | Tillisch (2017) | USA | Healthy | 40 | 100% | 28.9 (9.9) | No antibiotics month before inclusion | No probiotics month before inclusion | Fair |  |
|  | 36 | Kohn (2021) | Netherlands | Healthy | 58 | 100% | 21.5 (0.5) | No antibiotics (no period specified) |  | Fair |  |
| **Healthy population Children** | 34 | Gao (2019) | USA | Infants | 39 | 59% | 404 days (38.3) |  |  | Poor |  |
|  | 35 | Kelsey (2021) | USA | Newborns | 63 | 41% | 24.9 days (10.7) |  |  | Good |  |
| **Disease population Adults** | 43 | Dong (2020) | USA | (Obese) patients undergoing LSG | 14* | 100% | 37.4 (9.7) | No antibiotics month before inclusion | No probiotics month before inclusion | Fair | Pre- and post-surgery timepoints pooled in analyses |
|  | 41 | Strandwitz (2019) | USA | MDD | 23 | 65% | [19-65] (min-max) |  |  | Fair |  |
|  | 29, 30 | Wang (2019) / Zheng (2020) | China | ESRD / healthy | 28 / 19 | 54% / 37% | 43.9 (13.8) / 44.1 (10.0) | No antibiotics 2 weeks before inclusion | No probiotics (no period specified) | Poor |  |
|  | 37 | Li (2022) | China | BP / healthy | 44 / 37 | 50% / 54% | 22.0 (8.2) / 21.1 (1.9) | No antibiotics 4 weeks before inclusion | No pre-/probiotics 4 weeks before inclusion | Fair |  |
|  | 42 | Dong (2022) | USA | Obese / healthy | 81 / 216 | 72.8% / 64.4% | 32 [26-41.5] / 26 [21-34] | No antibiotics 3 months before inclusion |  | Fair |  |
|  | 40 | Labus (2019) | Sweden | IBS / Healthy | 65 / 21 | 71% | 33.3 (10.2) / 33.3 (9.6) | No antibiotics month before inclusion | No probiotics month before inclusion | Fair |  |
|  | 44 | Hong (2021) | China | (Obese) patients undergoing VSG | 16* | 67.0% | 28.6 (4.1) | No antibiotics 3 months before inclusion | No probiotics (incl. foods) 7 days before sample collection | Good |  |
|  | 39 | Jacobs (2021) | USA | IBS patients responding to CBT / not responding to CBT | 22 / 12 | 81.8% / 83.3% | 43.7 (13.1) / 48.0 (14.9) | No antibiotics 12 weeks before inclusion |  | Good |  |
|  | 38 | Ahluwalia (2016) | USA | Cirrhosis (with/without HE) | 74* | 41% | 54.2 (11.6) / 56.2 (13.5) | No antibiotics 6 weeks before inclusion |  | Poor | HE groups pooled in analyses |

\* the sample size refers to the total number of participants, as participants in different groups/data from different timepoints were pooled for the association analysis
**Abbreviations:** **BP:** bipolar disorder; **CBT:** cognitive behavioral therapy; **ESRD:** end-stage renal disease; **HE:** hepatic encephalopathy; **IBS:** irritable bowel syndrome; **LSG:** laparoscopic sleeve gastrectomy; **MDD:** major depressive disorder; **VSG:** vertical sleeve gastrectomy

### 4.2 Methodological characteristics (***table 2***)

An overview of the methods and indices used to quantify and analyze the gut microbiota and functional and structural brain connectivity is provided in **Figure 2.**

**Figure 2.**
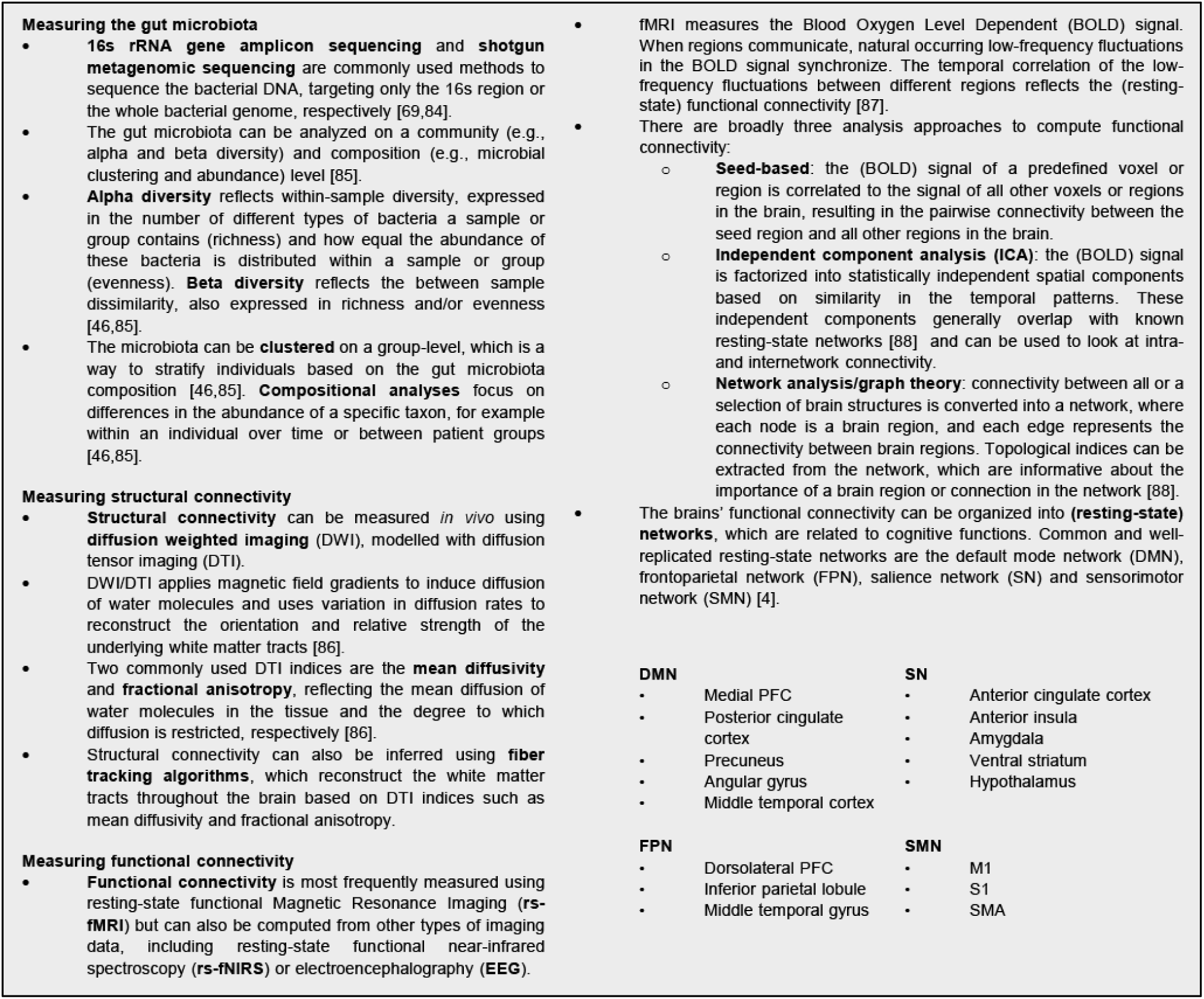
Overview of commonly used techniques and indices to investigate the gut microbiota and functional and structural brain connectivity.

#### 4.2.1 Microbiota quantification

The included studies employed various sequencing workflows to estimate the gut microbiota composition. Three studies performed shotgun metagenomic sequencing [35,37,44], while all other studies performed 16s rRNA gene sequencing.

##### Microbial Diversity

Six out of sixteen studies tested the association between alpha diversity/richness and connectivity [26,27,29–32,34,35], and one out of sixteen studies assessed beta diversity [34].

##### Microbial Composition

Three studies performed microbiota-based clustering [26,27,29,30,33], and all but two studies assessed the microbial abundance. Additional methodological information regarding sample collection, and data processing is presented in **Table 2**.

**Table 2.**
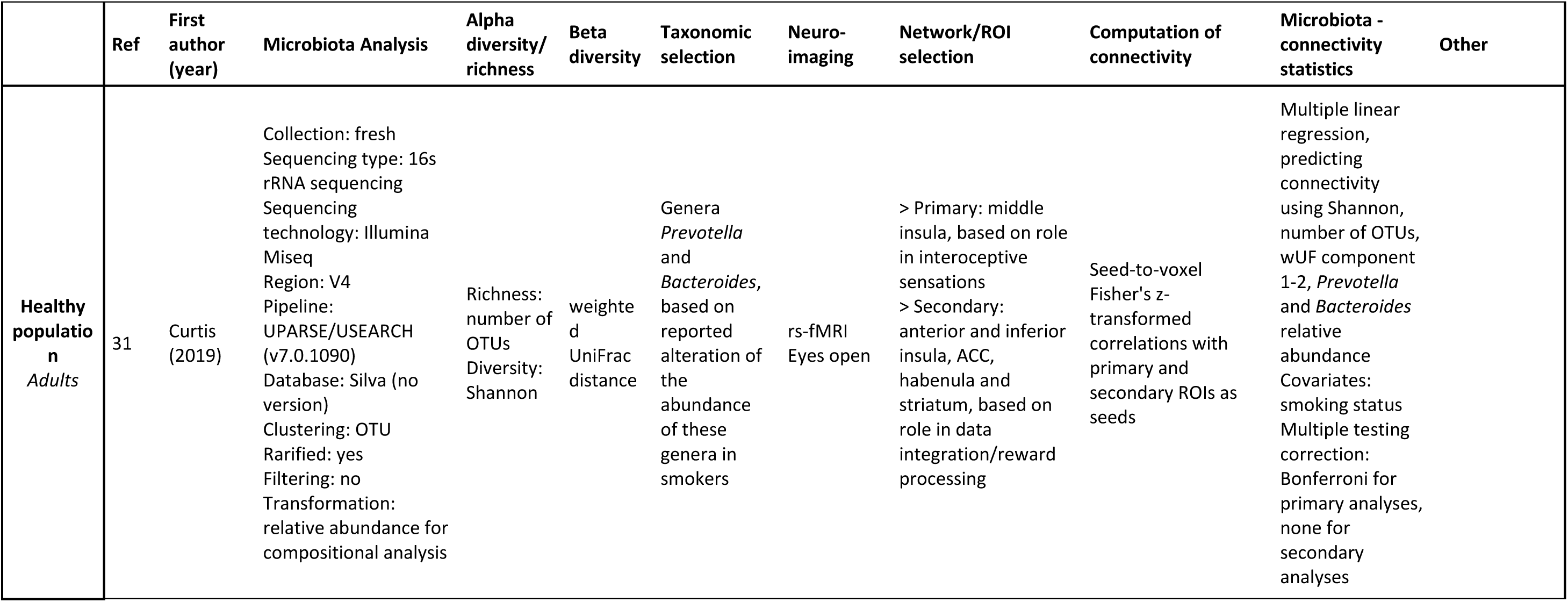
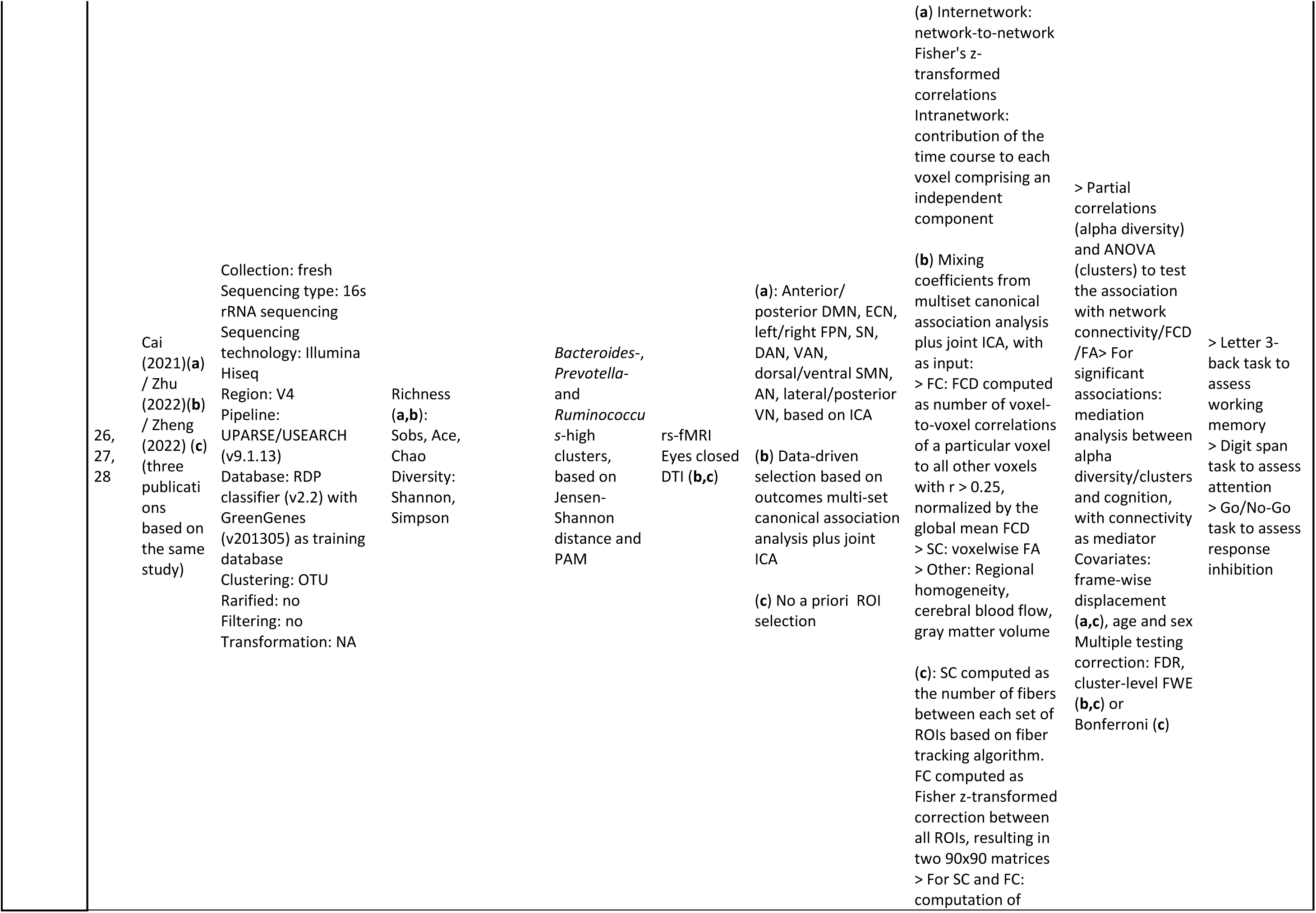
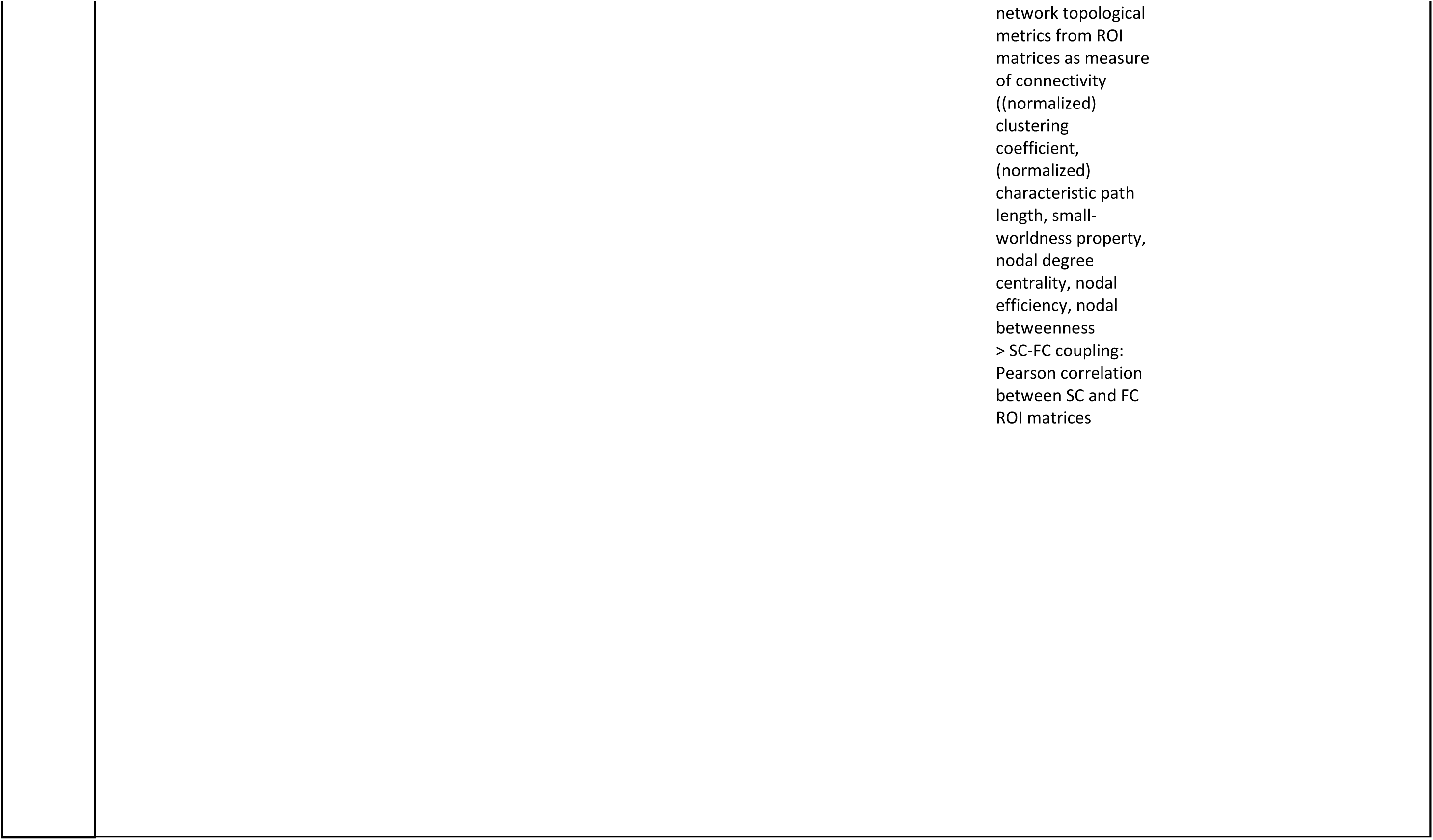
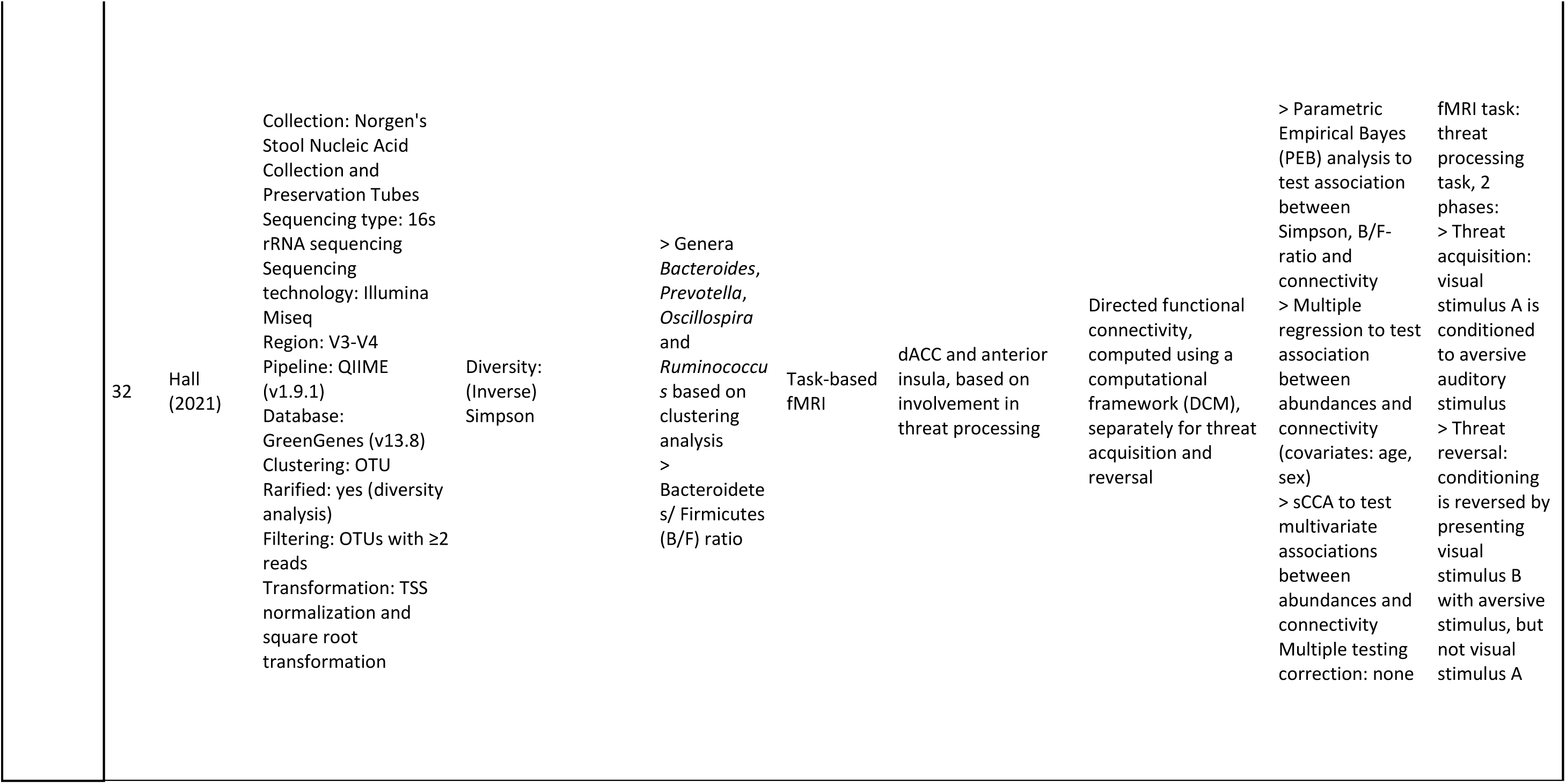
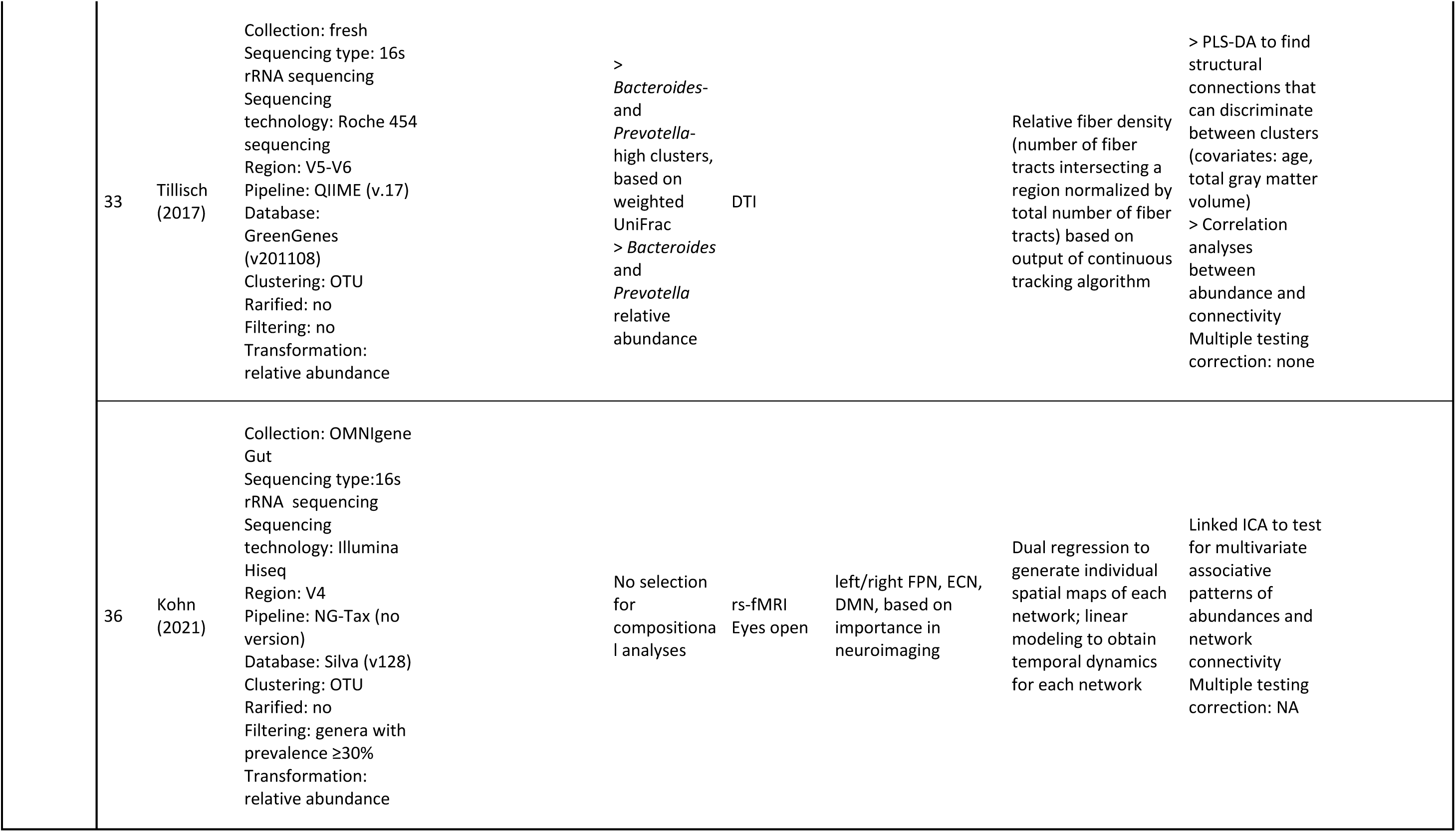
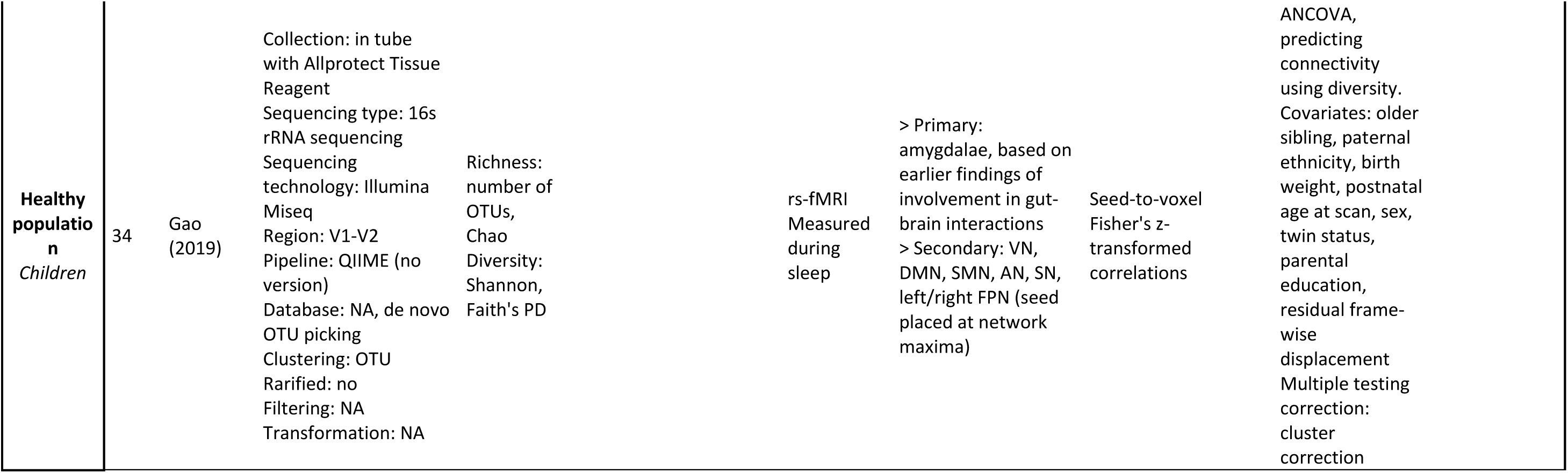
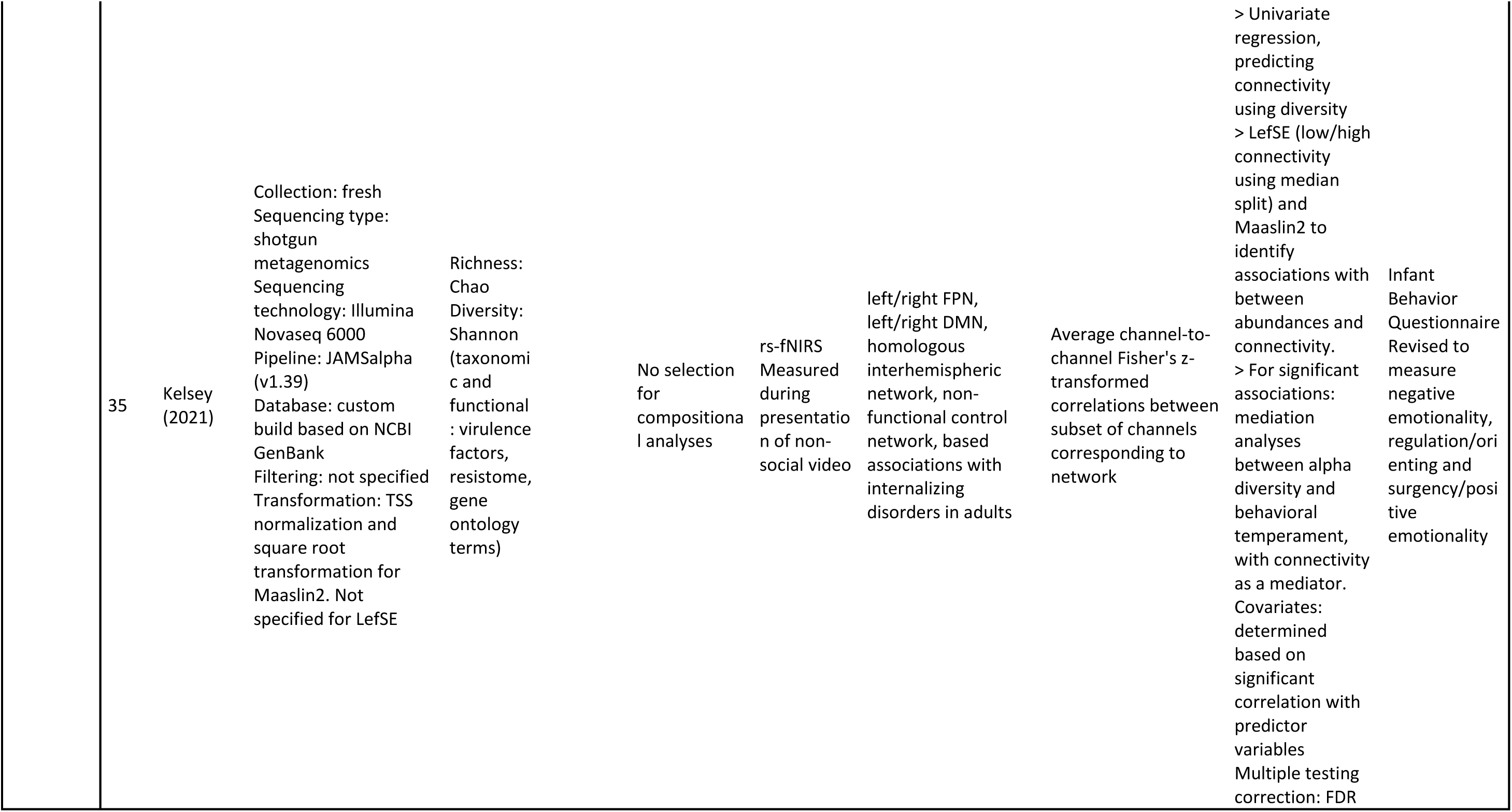
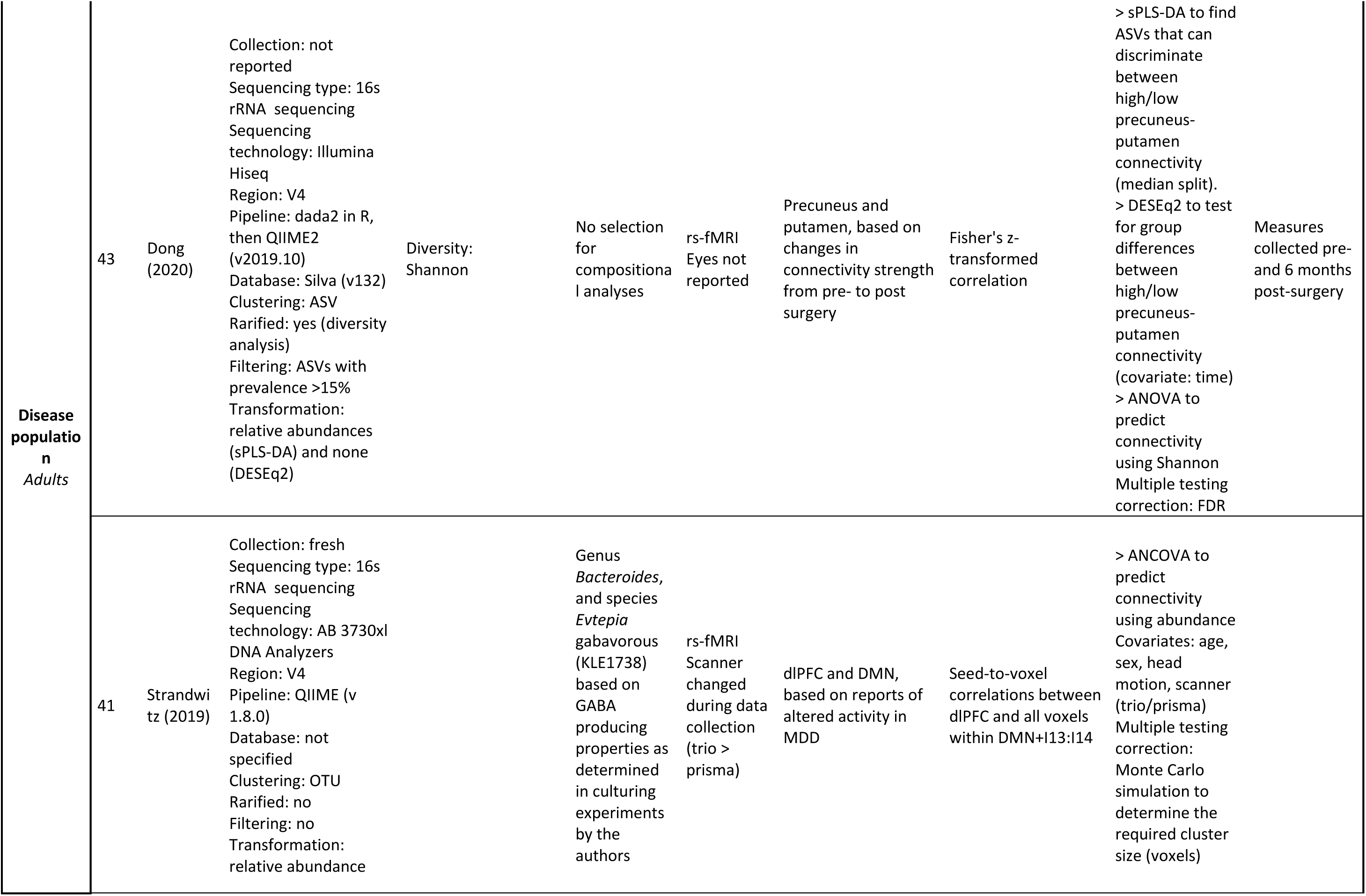
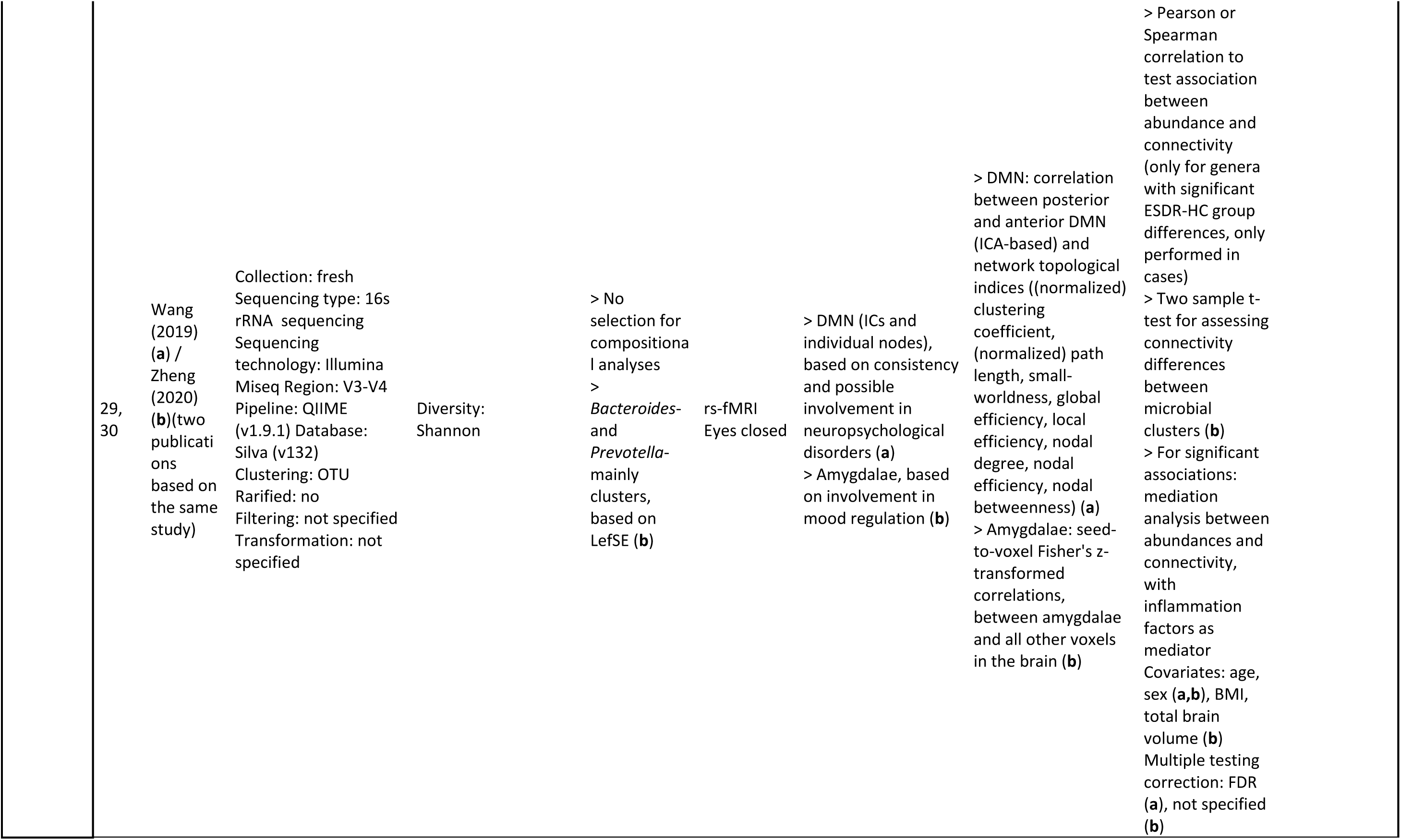
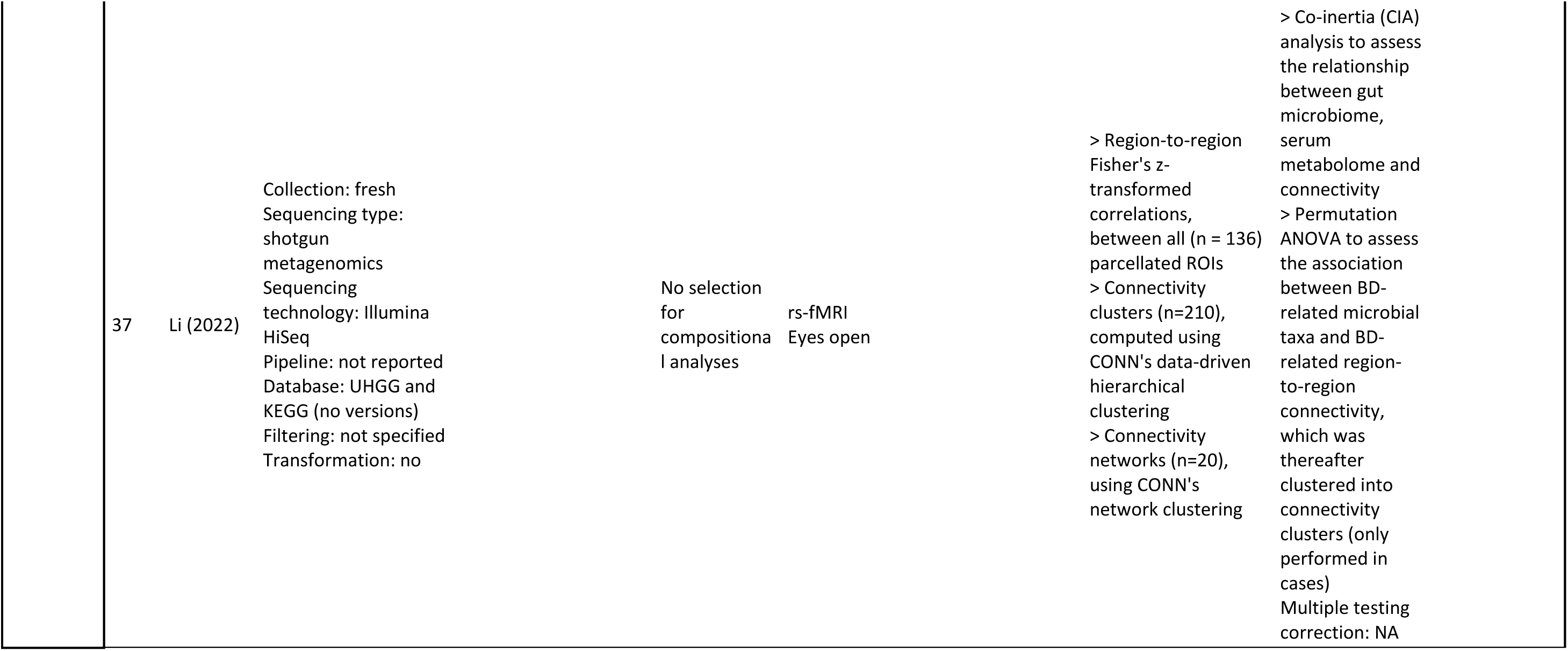
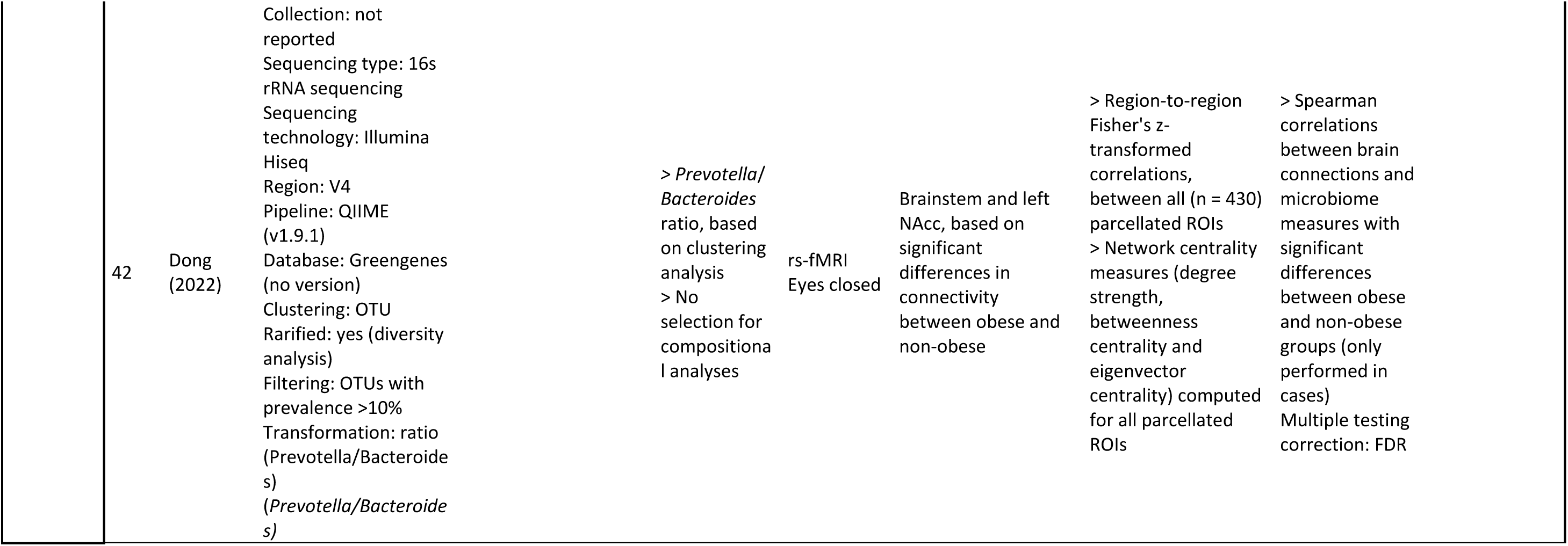
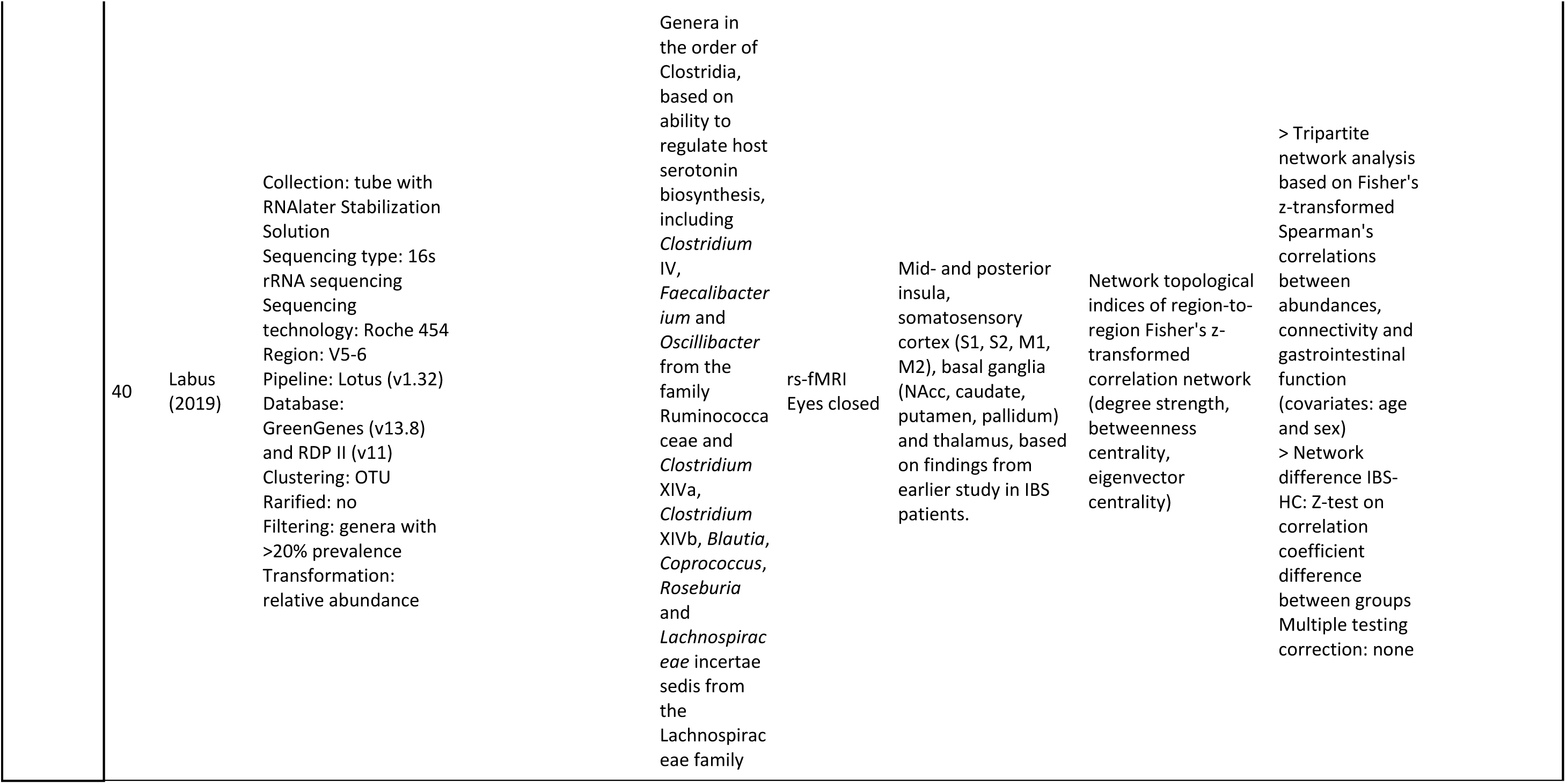
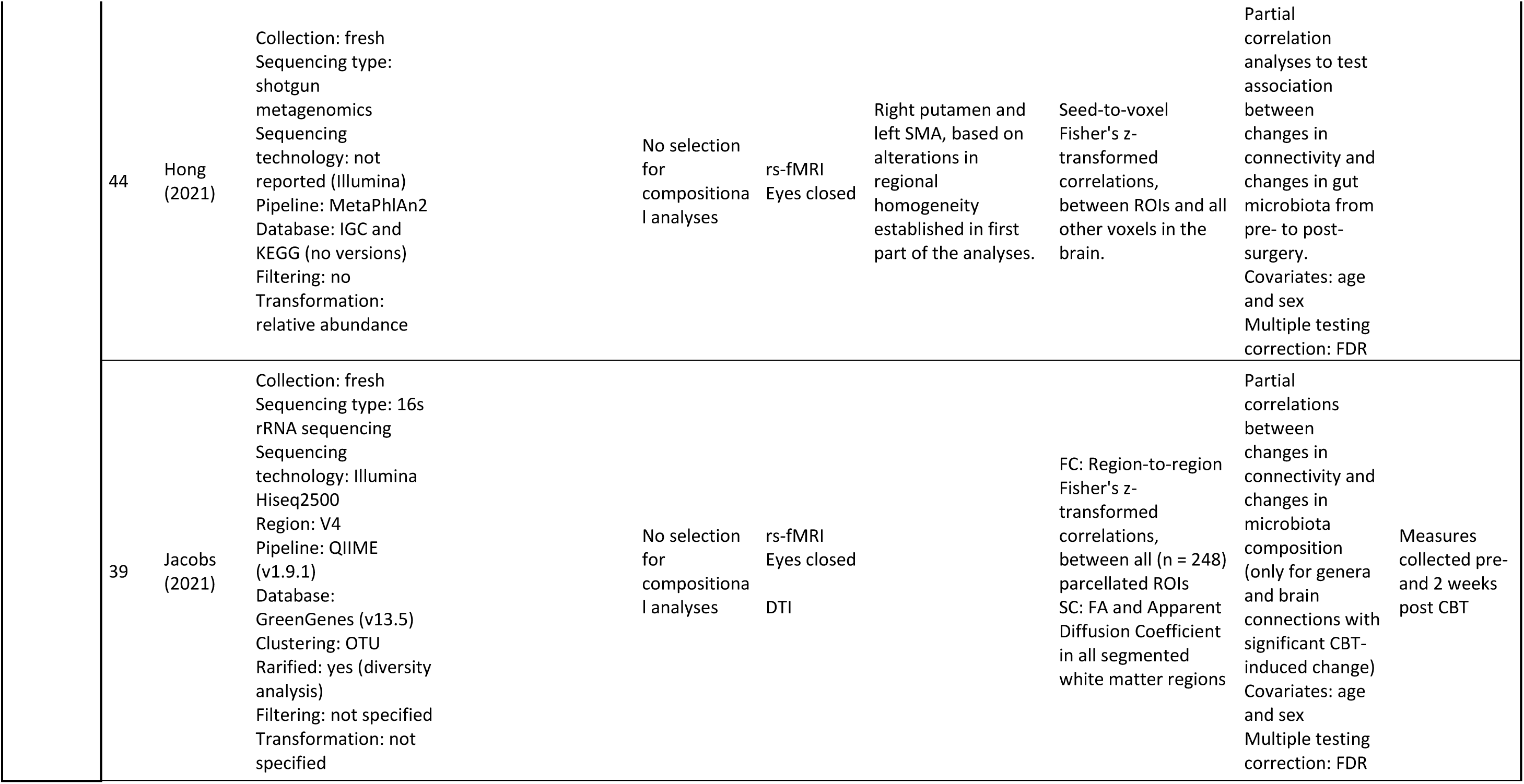
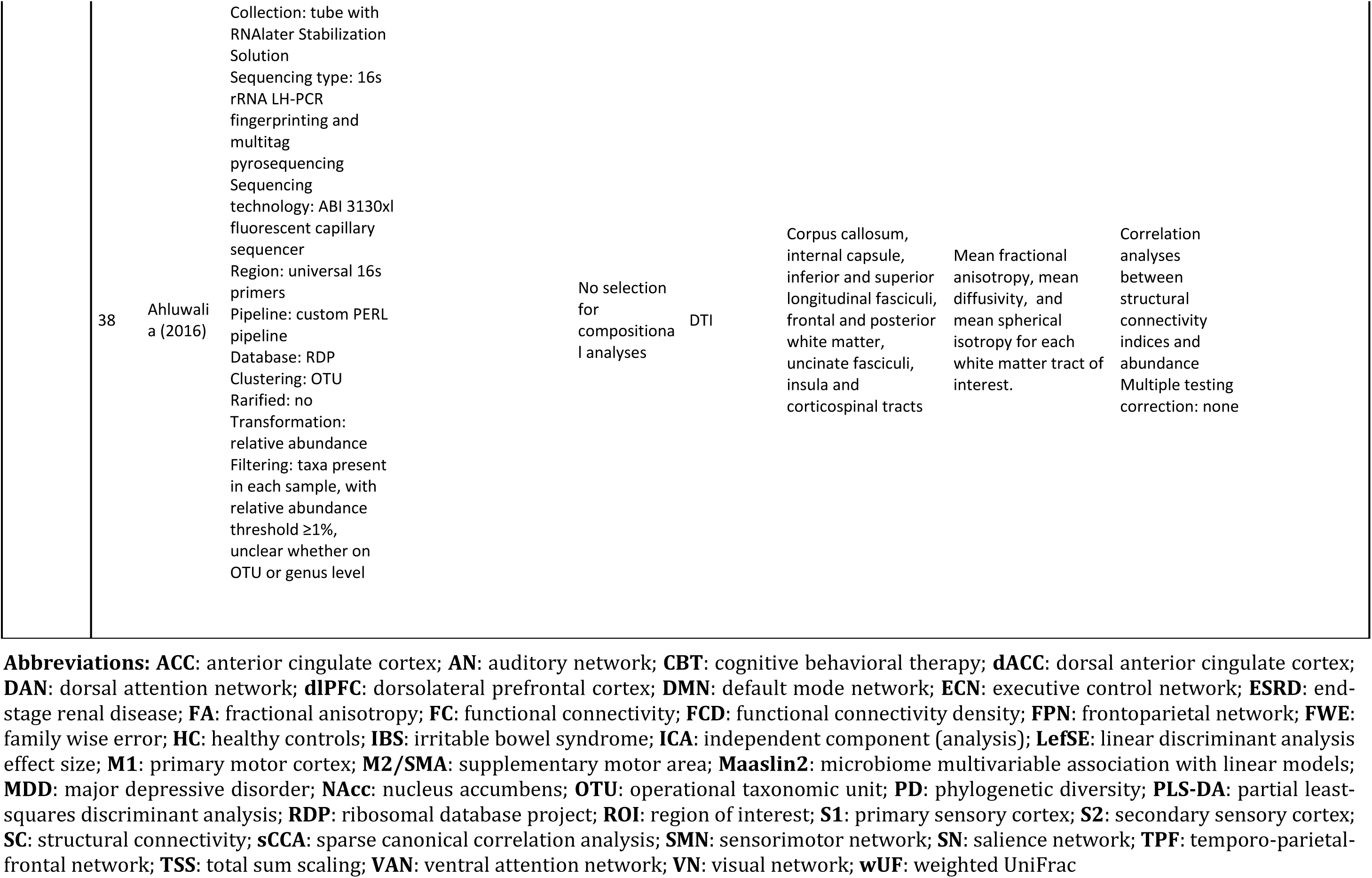
Overview of methodological characteristics

#### 4.2.2 Connectivity quantification

##### Functional connectivity assessment

Fourteen out of sixteen studies assessed functional connectivity, of which twelve used resting-state functional magnetic resonance imaging (rs-fMRI), one used task-based fMRI [32], and one used resting-state functional near infrared spectroscopy (fNIRS) [35].

##### Structural connectivity assessment

Four out of fourteen studies assessed structural connectivity using diffusion tensor imaging (DTI) [27,33,38,39]. Additional information about the metrics used to compute connectivity, and the *a priori* selection of brain regions, networks or white matter tracts for each study is provided in in **Table 2**.

#### 4.2.3 Statistical analysis on microbiota-connectivity association

The included studies employed various statistical methods to explore the association between microbiota and brain connectivity, including linear regression/ANCOVA [31,32,34,35,45], (permutation) ANOVA [26,27,37,43], (partial) correlation [26–30,33,38,39,42,44] and tripartite network analysis, based on correlations [40]. Three studies used packages for differential abundance testing, including sparse partial least squares discriminant analysis (sPLS-DA) [33, 43], linear discriminant analysis effect size (LefSE) [35], microbiome multivariable association with linear models (MaAsLin2) [35] and differential gene expression analysis based on the negative binomial distribution (DESeq2) [43]. Finally, linked ICA [36], spatial canonical correlation analysis (sCCA) [32] and Parametric Empirical Bayes (PEB) [32] analysis were employed by one study each.

### 4.3 Synthesized results (***table 3***)

First, studies investigating healthy participants are discussed, covering studies in adults first, followed by studies in children. Second, studies in a disease population are discussed. In each section, results on microbial diversity will be discussed first, followed by results on microbial composition (microbial clustering and abundance).

**Table 3.**
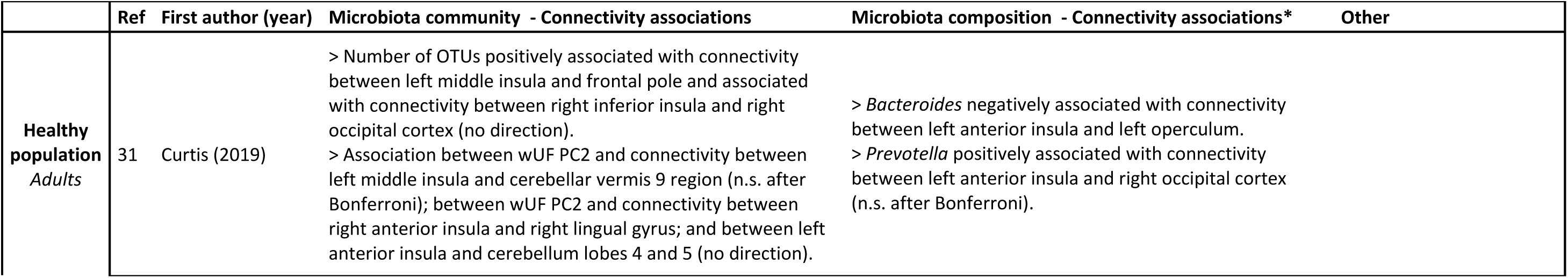
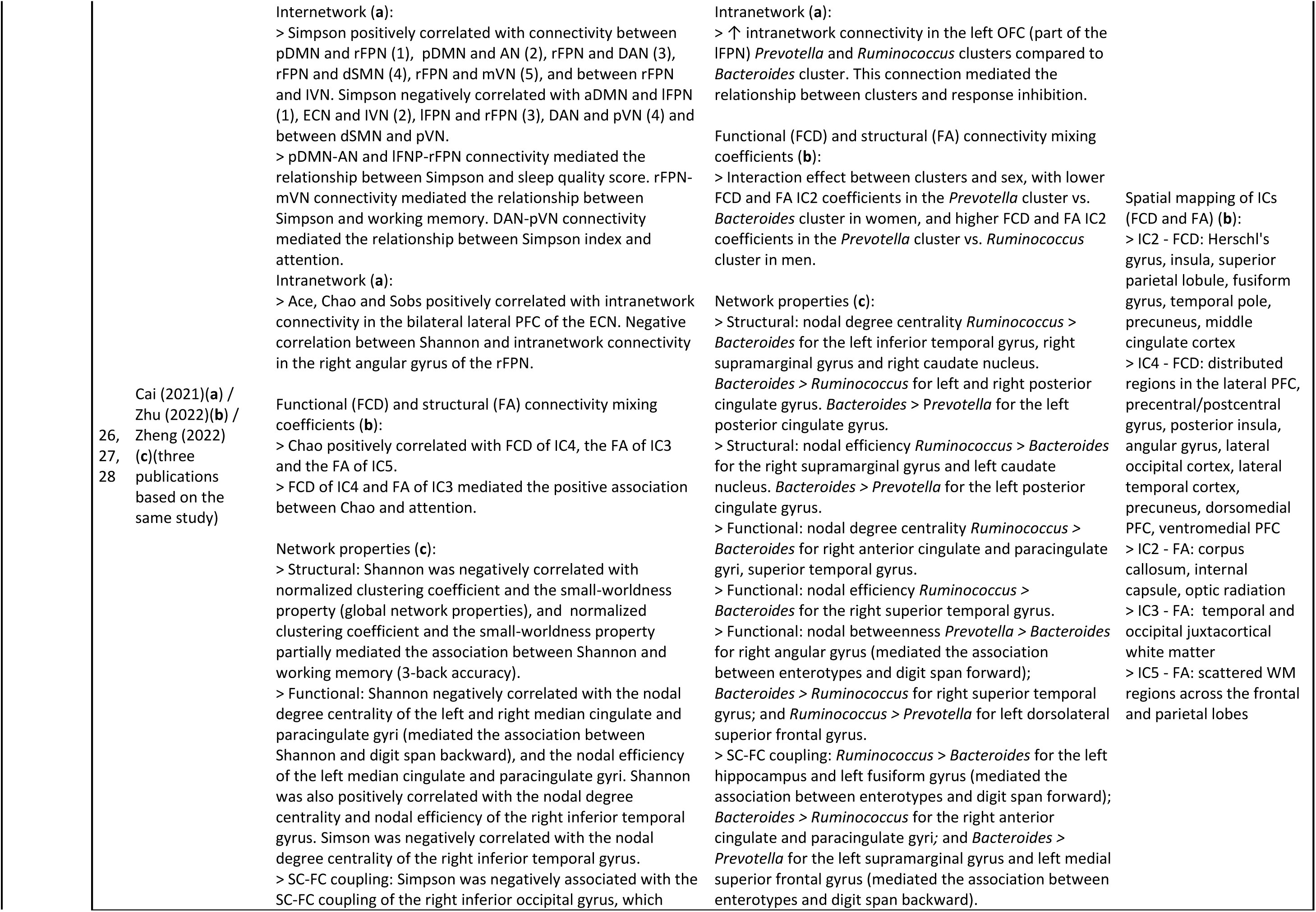
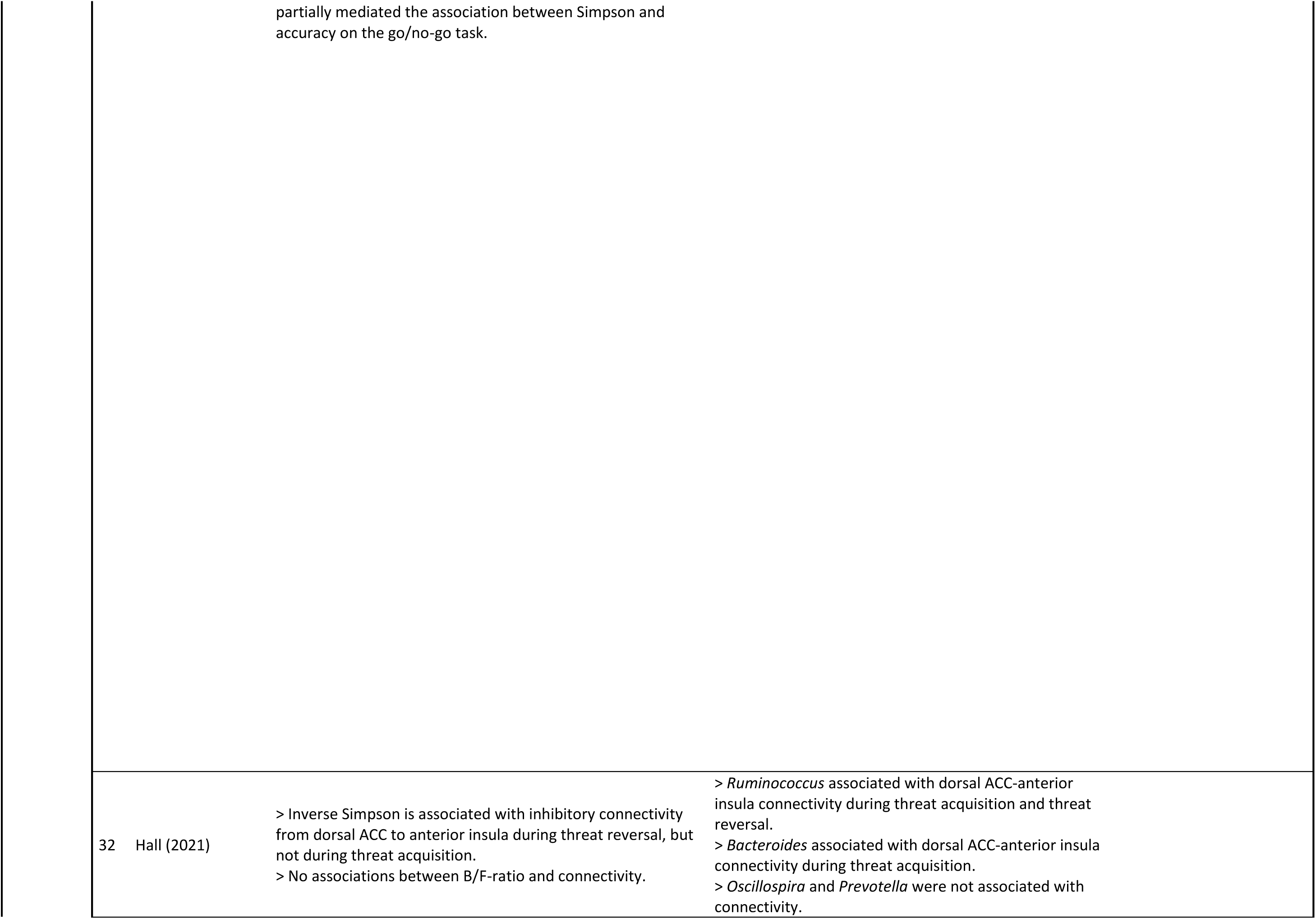
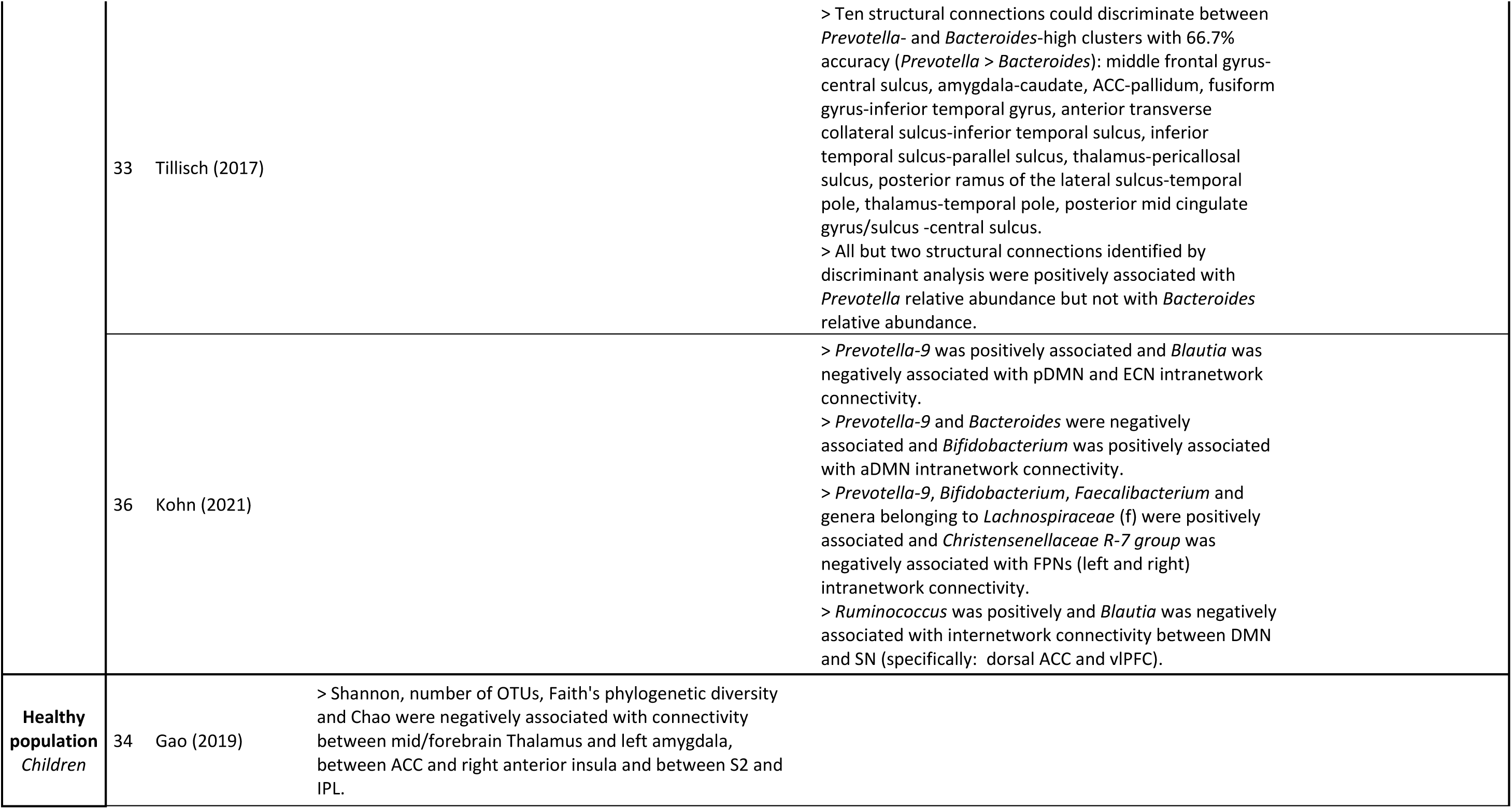
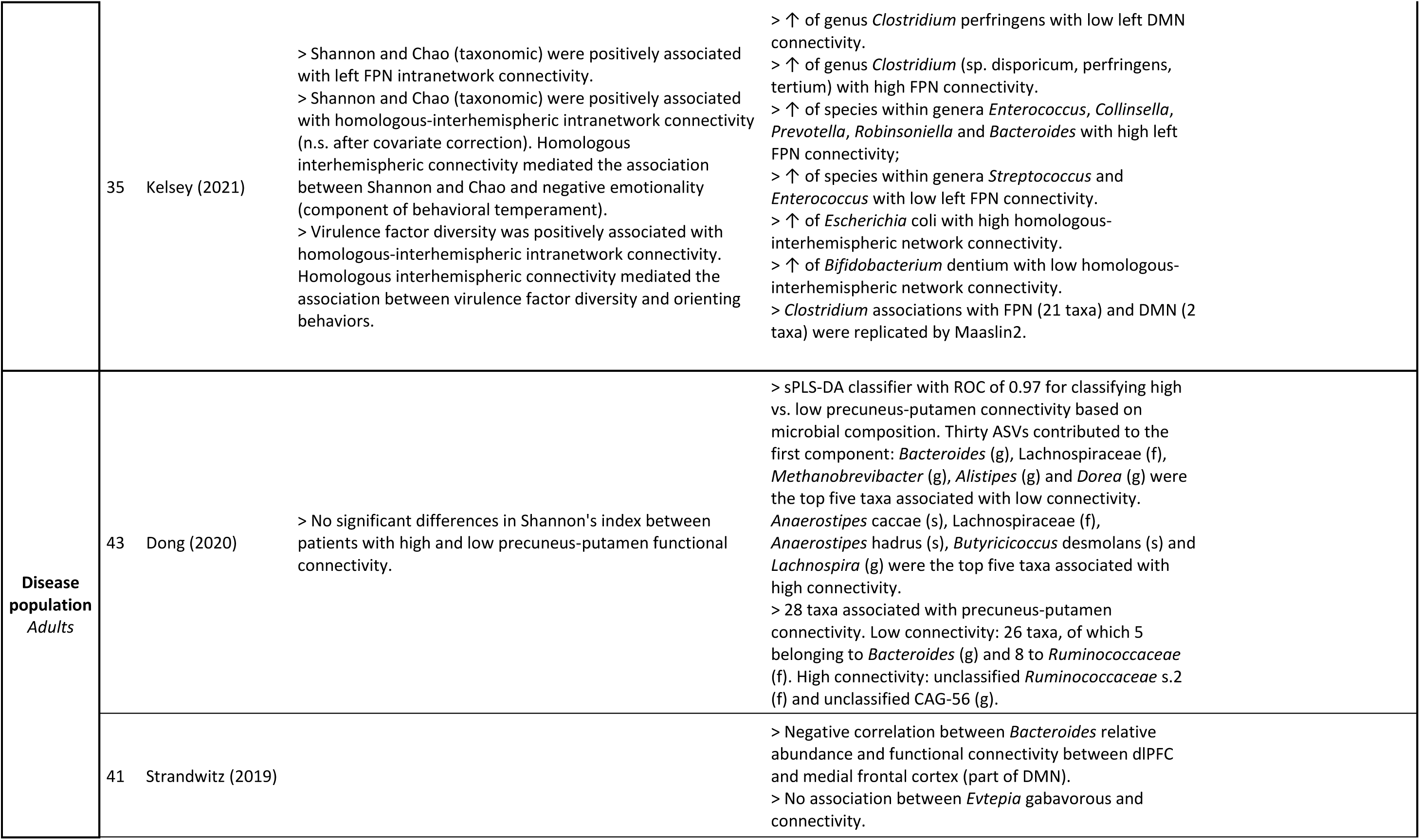
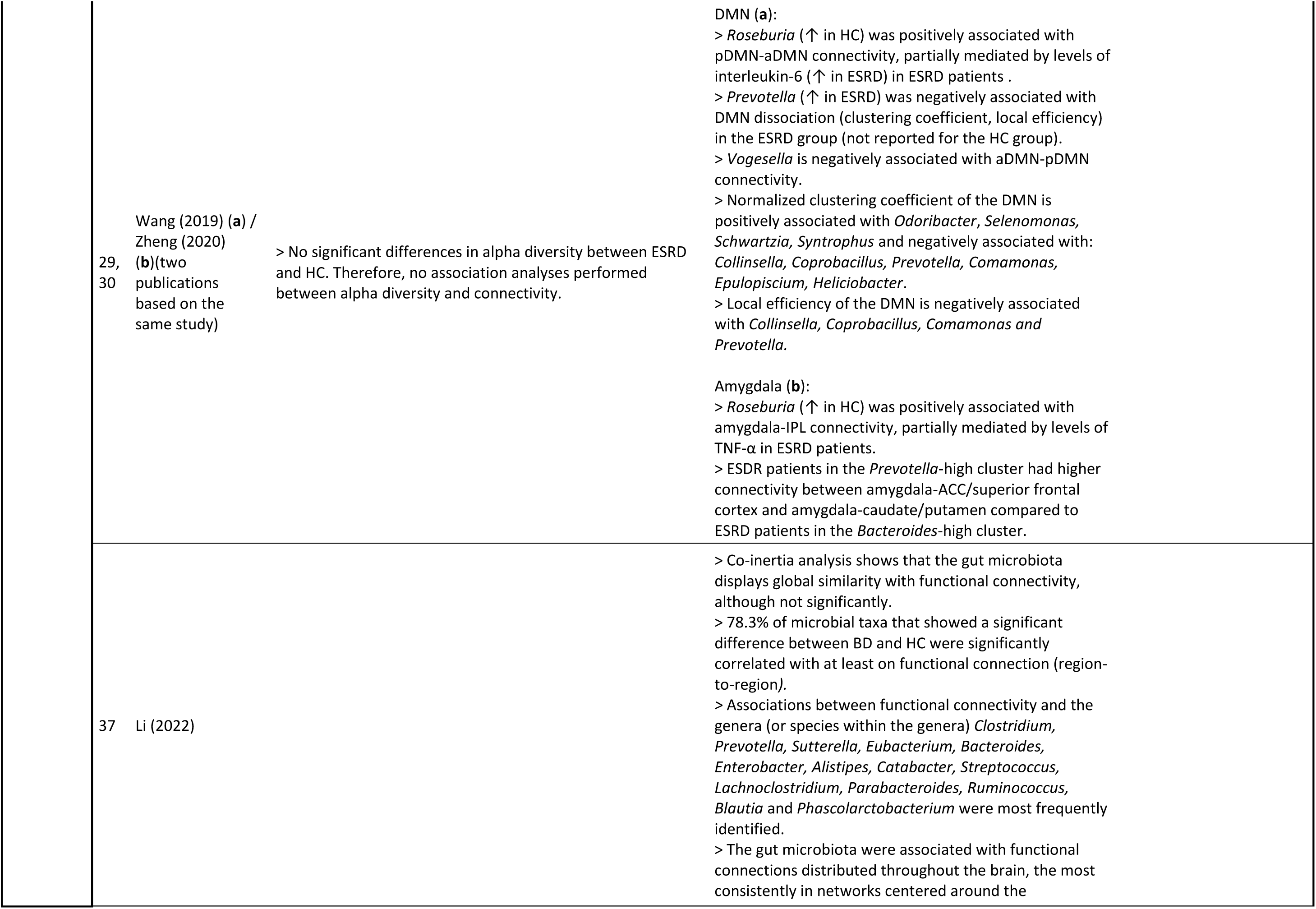
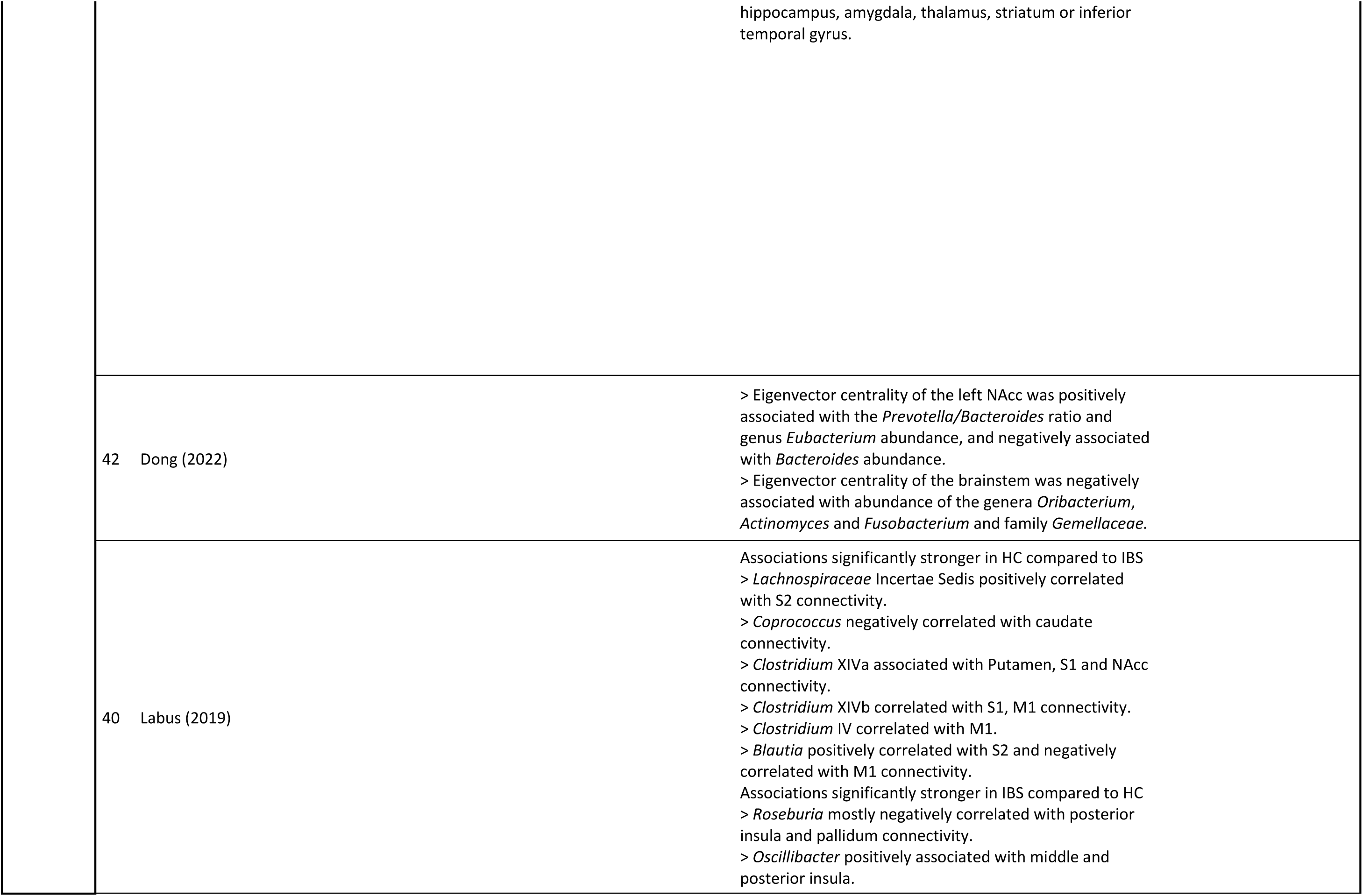
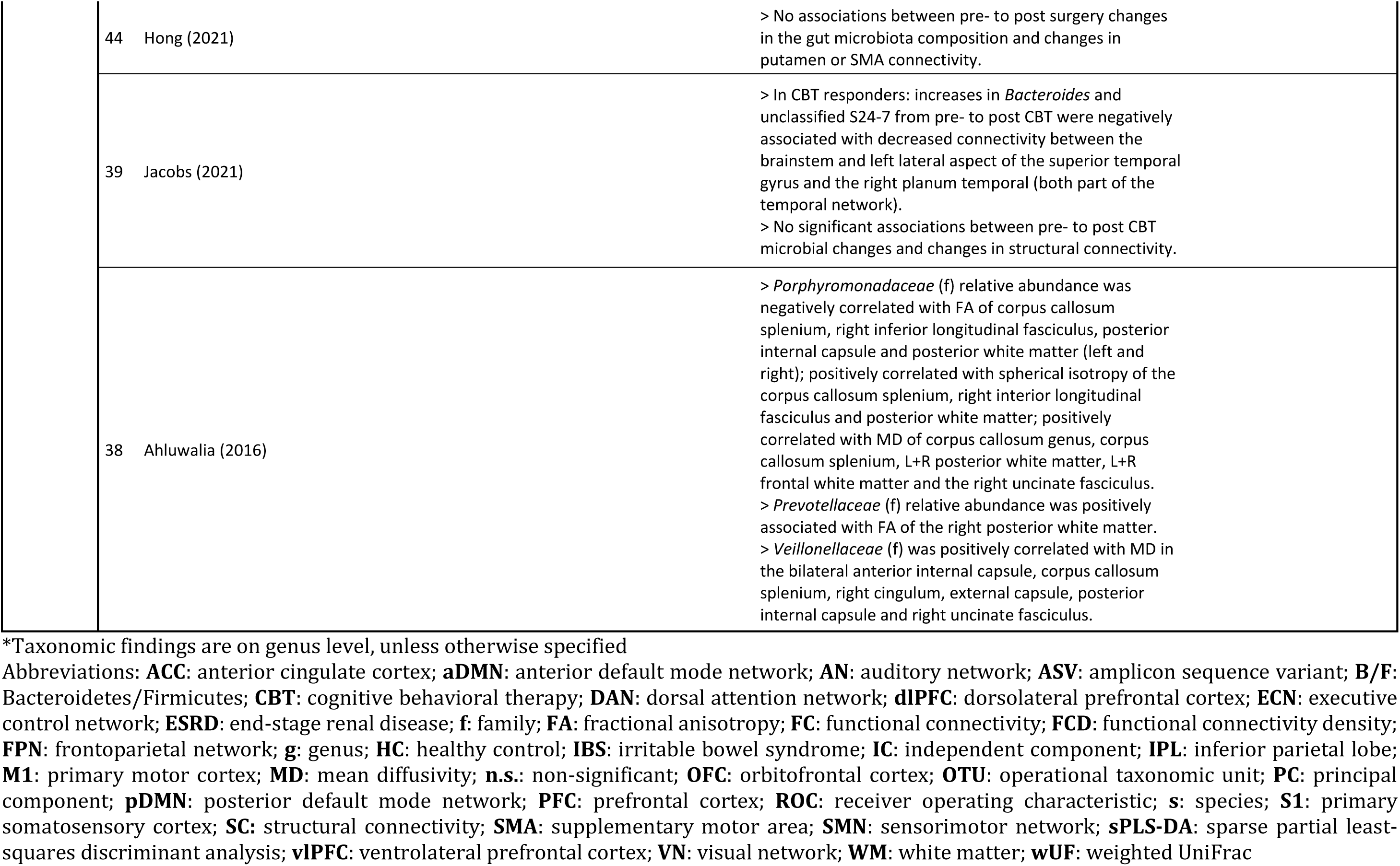
Overview of synthesized study findings

#### 4.3.1 Healthy population: adults

##### Microbial diversity

Three studies have assessed functional connectivity and microbial alpha and/or beta diversity in a sample of healthy adults. Curtis and colleagues [31] assessed the association between seed-based functional connectivity of the insula and alpha and beta diversity, and reported an association between microbial richness (number of OTUs) and the functional connectivity between the insula and occipital cortex. The direction of the association was not reported. Regarding beta diversity, associations between weighted UniFrac distance and the connectivity between the insula and the operculum (non-significant after Bonferroni correction), lingual gyrus and cerebellum were reported.

A study conducted by Cai and colleagues, Zhu and colleagues, and Zhang and colleagues [26–28] yielded three publications. In the first publication [26] the authors reported a positive association between microbial richness (Sobs, Ace, Chao) and intra-network connectivity of the executive control network and between alpha diversity (Shannon) and intra-network connectivity of the FPN. Moreover, alpha diversity (Simpson) was associated with inter-network connectivity between networks related to executive control, default mode and sensorimotor systems. In the second publication [27], the authors associated multimodal independent components, based on among others functional connectivity density and fractional anisotropy, with richness and diversity. Using this approach, microbial richness (Chao) was found associated with numerous functional and structural connections distributed throughout the brain. The third publication [28] focused on functional and structural network organization, identifying associations between alpha diversity (Shannon, Simpson), global network properties of the structural network, and regional network properties of the functional network, mainly within the cingulate and temporal cortex. Additionally, there were associations between alpha diversity (Simpson) and the coupling between structural and functional connectivity of occipital regions. Finally, Hall and colleagues [32] investigated the association between alpha diversity (Simpson) and the directed functional connectivity between the dorsal anterior cingulate cortex (dACC) and anterior insula during threat processing (threat acquisition and reversal), and revealed that there was a positive association between alpha diversity and the inhibitory connectivity from the dACC to the anterior insula connectivity during threat reversal, but not during threat acquisition.

##### Microbial composition: clustering

Two studies performed microbiota-based clustering to associate the gut microbiota with functional and/or structural brain connectivity in healthy adults. The three publications based on the study by Cai and colleagues, Zhu and colleagues and Zhang and colleagues [26–28] describe three microbiota-based clusters characterized by high *Prevotella*, high *Bacteroides*, or high *Ruminococcus* abundance. In the first publication [26], associating these clusters to fourteen ICA-based resting-state networks showed that the participants within the *Prevotella-* and *Ruminococcus-*clusters had higher intra-network connectivity of the left FPN compared to participants in the *Bacteroides*-cluster. In the second publication [27], the authors used a multimodal independent components and identified sex-dependent differences between microbial clusters in functional and structural connectivity strength. In the third publication [28], differences between clusters were reported in both functional and structural network organization, both in terms of global and regional network properties. Moreover, there were differences between clusters in the coupling between functional and structural connectivity in a distributed set of brain regions.

Finally, Tillisch and colleagues [33] identified two clusters, one characterized by high *Prevotella* abundance (containing seven participants), and one characterized by *Bacteroides* abundance (containing 33 participants), and investigated which structural connections could best discriminate between the two clusters. Discriminant analysis revealed ten structural connections, distributed throughout the brain, whose strength could discriminate between the clusters with 66.7% accuracy.

##### Microbial composition: abundance

Four studies assessed the association between microbial abundance and functional (three studies) or structural (one study) brain connectivity in healthy adults. Curtis and colleagues [31], assessing the genera *Prevotella* and *Bacteroides*, reported a negative association between *Bacteroides* relative abundance and the functional connectivity between the anterior insula and the operculum, and a positive association between *Prevotella* relative abundance and the functional connectivity between the anterior insula and the occipital cortex, the latter being non-significant after Bonferroni correction.

Hall and colleagues [32] assessed the association between the abundances of *Bacteroides*, *Prevotella*, *Oscillospira*, and *Ruminococcus* and directed threat-related functional connectivity, revealing an association between *Bacteroides* abundance and dACC-anterior insula connectivity during threat acquisition. Additionally, *Ruminococcus* abundance was associated with dACC-anterior insula connectivity during threat acquisition and threat reversal. For threat reversal, this association could be attributed to a positive association between *Rumicococcus* abundance and connectivity from the anterior insula to the dACC.

Kohn and colleagues [36] performed multivariate linked ICA and identified four independent components with contribution from both the gut microbiota and functional network connectivity. First, *Prevotella* abundance was positively and *Blautia* abundance was negatively associated with the intra-network connectivity of the posterior DMN and executive control network. Second, *Prevotella* and *Bacteroides* abundance were negatively, and *Bifidobacterium* abundance was positively associated with intra-network connectivity of the anterior DMN. Third, the abundance of *Bifidobacterium*, *Faecalibacterium* and genera belonging to the *Lachnospiracaceae* family were positively and *Christensenellacea_R-7* was negatively associated with the intra-network connectivity of the FPNs. Fourth, *Ruminococcus* abundance was positively and *Blautia* abundance was negatively associated with the internetwork connectivity between the DMN and SN.

Finally, Tillisch and colleagues [33] assessed the association between *Prevotella* and *Bacteroides* relative abundance and the ten structural connections with the highest explanatory value in differentiating between *Prevotella* and *Bacteroides* microbiota clusters, as discussed above. *Prevotella* relative abundance was associated with all but two of the ten structural connections, while *Bacteroides* relative abundance was not associated with any of the connections.

#### 4.3.2 Healthy population: children

##### Microbial diversity

Two studies assessed functional brain connectivity and microbial alpha diversity in a sample of healthy children. Gao and colleagues [34] investigated one-year-old infants and reported a negative association between microbial richness (number of OTUs, Chao) and diversity (Shannon, Faith’s PD) and the connectivity between the amygdala and thalamic regions, and between the anterior cingulate cortex and anterior insula. Additionally, there was a positive association between alpha diversity/richness and the connectivity between the supplementary motor area and the inferior parietal lobule.

Kelsey and colleagues [35] assessed taxonomic and functional alpha diversity in one-month-old infants, revealing a positive association between taxonomic alpha diversity/richness (Chao, Shannon) and frontoparietal and homologous-interhemispheric intranetwork connectivity. Additionally, there was a positive association between virulence factor diversity (i.e., diversity of bacteria producing molecules associated with disease) and intranetwork connectivity of the homologous-interhemispheric network.

##### Microbial composition: abundance

Kelsey and colleagues [35] explored the association between functional brain connectivity and gut microbiota composition in one-month-old infants, revealing enrichment of species within the genus *Clostridium* in infants with high frontoparietal and low default mode intranetwork connectivity. Moreover, high frontoparietal intranetwork connectivity was characterized by enrichment of species *Enterococcus* faecalis*, Collinsella* (unclassified species), *Prevotella* copri, *Robinsoniella* peoriensis and *Bacteroides* caccae, whereas *Enterococcus* (unclassified species) and *Streptococcus* salivarius were enriched in low left frontoparietal intranetwork connectivity. Finally, high homologous-interhemispheric intranetwork connectivity was characterized by enriched *Escherichia* coli, and low homologous-interhemispheric intranetwork connectivity by enriched *Bifidobacterium* dentium.

#### 4.3.3 Disease population: adults

##### Microbial diversity

The association between alpha diversity and (functional) brain connectivity in a disease population was only assessed by one study. Dong and colleagues [43] examined the link between alpha diversity (Shannon) and functional connectivity between the precuneus and putamen in obese patients undergoing a laparoscopic sleeve gastrectomy. Data pooled from before and after surgery showed no differences in alpha diversity between patients with high and low precuneus-putamen connectivity.

##### Microbial composition: abundance

Nine studies assessed brain connectivity and microbial composition in a disease population, of which two were case-only studies with one timepoint, four were case-controlled with a group of healthy adults, and three were case-only using a longitudinal design. Eight out of nine studies assessed functional connectivity and two studies assessed structural connectivity. Strandwitz and colleagues [41] investigated the association between relative abundance of the genus *Bacteroides* and the functional connectivity between the left dorsolateral prefrontal cortex (dlPFC) and the DMN in patients with major depressive disorder, and reported that *Bacteroides* relative abundance was negatively correlated with dlPFC-DMN connectivity.

A study performed by Wang and colleagues and Zheng and colleagues [29, 30] in patients with end-stage renal disease yielded two publications. In the first publication [29] the authors assessed the association between intranetwork DMN connectivity and microbial composition, and reported a positive association between *Roseburia* relative abundance and connectivity between the anterior and posterior DMN. This association was partially mediated by levels of the pro-inflammatory cytokine interleukin-6. The relative abundances of *Colinsella, Coprobacillus, Comamonas, Epulopiscium, Heliciobacter, Odoribacter*, *Prevotella, Schwartzia, Selenomonas, Syntrophus* and *Vogesella*, were also associated with DMN network organization (clustering coefficient and local efficiency), but these associations were not mediated by inflammation markers. In the second publication [30], the authors focused on the association between microbial composition and seed-based amygdala functional connectivity, and identified an association between *Roseburia* absolute abundance and the connectivity between the amygdala and the inferior parietal lobule. This association was partially mediated by levels of the pro-inflammatory cytokine tumor necrosis factor alpha.

Li and colleagues [37] assessed the gut microbiota composition and functional connectivity in patients with bipolar disorder. A total of 78% of microbial taxa that were differentially abundant in cases compared to controls were also associated with at least one functional connection in the brain. The genera *Clostridium*, *Prevotella* and *Suterella* were most frequently reported, together with functional networks centered around the hippocampus, amygdala, thalamus, striatum, and inferior temporal gyrus.

Dong and colleagues [42] investigated the gut microbiota composition and functional brain network properties in individuals with obesity. The authors reported associations between the eigenvector centrality, a metric reflecting the influence of one region in a network, of the nucleus accumbens and the ratio between *Prevotella* and *Bacteroides* abundance, as well as *Eubacterium* abundance. Additionally, the eigenvector centrality of the brainstem was associated with abundance of the genera *Oribacterium*, *Actinomyces* and *Fusobacterium*.

Labus and colleagues [40] performed a case-control study to explore the association between relative abundance of genera in the order *Clostridia* and resting-state functional connectivity of sensorimotor brain regions in patients with irritable bowel syndrome (IBS) and healthy adults. Using tripartite network analysis, significant differences in the associations between sensorimotor brain regions and *Clostridium* (*XIVa* and *XIVb)*, *Coprococcus,* and an unclassified Lachnospiraceae genus were identified between cases and controls: associations were observed in healthy adults but were largely absent in patients with IBS. One exception is the *Roseburia* genus: in patients with IBS, there were multiple, mostly negative, associations between this genus and connectivity, whereas such associations were absent in healthy adults.

Dong and colleagues [43] examined the association between gut microbiota composition and the connectivity between the precuneus and putamen in obese patients undergoing laparoscopic sleeve gastrectomy. Discriminant analysis showed that the microbial composition could discriminate between participants with a high and low precuneus-putamen connectivity with an ROC of 0.97. *Bacteroides, Methanobrevibacter, Alistipes* and *Dorea* were enriched in participants with high precuneus-putamen connectivity, and *Anerostipes*, *Lachnospira* and *Butyricococcus* were enriched in participants with a low precuneus-putamen connectivity.

Hong and colleagues [44] performed a longitudinal study to examine the association between changes in gut microbiota composition and changes in seed-based connectivity of the putamen and supplementary motor area from pre-to post vertical sleeve gastrectomy in obese patients. The surgery did induce changes in functional connectivity and microbial composition, but the changes were not associated.

Jacobs and colleagues [39] performed a longitudinal study to investigate how cognitive behavioral therapy (CBT) affected the gut microbiota composition and functional and structural connectivity in responding and non-responding patients with irritable bowel syndrome, and how such CBT-induced changes were associated. The authors reported increases in *Bacteroides* and unclassified S24-7 relative abundance in CBT responders compared to non-responders. Those changes were furthermore associated with decreased functional connectivity between the brainstem and regions within the superior temporal gyrus. CBT also induced changes in structural connectivity, but those were not associated with changes in the microbial composition.

Finally, Ahluwalia and colleagues [38] investigated the association between microbial composition and structural connectivity strength of the brain’s major white matter tracts in patients with cirrhosis, revealing associations between the abundance of the bacterial families *Porphyromonadaeae*, *Prevotellaceae* and *Veillonellaceae* and structural connections distributed throughout the brain.

### 4.4 Quality assessment

A majority of the studies were rated as ‘Fair’ (n=9), four studies were rated as ‘Poor’ and only three studies were rated as ‘Good’. The primary source of bias was related to incomplete reporting of the methods, including a lack of detail in the description of the recruitment procedure, microbiota data handling and description of the used statistical methods. Additionally, about a third of the studies did not correct for the effects of key confounders. The quality assessment and explanatory notes per study are shown in **Table S6**.

## 5 Discussion

### 5.1 Summary

A qualitative systematic synthesis of the available study findings shows associations between the gut microbiota and brain connectivity (see **Fig. 3** and **Table S1-2** for an overview). The genera *Bacteroides*, reported in nine out of eleven studies, and *Prevotella,* reported in six out of ten studies, were most consistently associated with brain connectivity. Additionally, genera within the order *Clostridiales* were also repeatedly reported in association with brain connectivity. Within this order, the genus *Ruminococcus* was most consistently reported (in four out of seven studies), followed by *Blautia* (in three out of five). These genera were associated with regions distributed throughout the brain, both on a region-to-region and network level, without a clear pattern emerging. That is, microbial genera were not associated with brain connections in a specific manner. (**Fig. 3b, Table S1**). Furthermore, out of six studies assessing microbial richness or diversity, five reported associations with brain connectivity on at least one richness/diversity index. Again, associations were found with the connectivity of a set of distributed brain regions and networks (**Fig. 3a, Table S1**). In terms of functional brain connectivity, DMN connectivity and FPN connectivity (reported in six out of six and four out of four studies, respectively) were consistently associated with the gut microbiota. Connectivity of the SN, particularly connectivity of the insula and (dorsal) anterior cingulate cortex (reported in six out of eight studies), showed high consistency in its association with the gut microbiota (**Fig. 3, Table S2**). These networks were exhibiting associations with both microbial diversity and composition. Most studies have only assessed functional connectivity, but the gut microbiota also revealed associations with the brains structural wiring. From this, however, no clear pattern emerged due to high divergence in the way structural connectivity was quantified. Finally, a majority of the studies differed in their target population, focusing on a range of different diseases. Considering the large methodological differences between studies as well as lack of direct comparisons between the case and control participants, is not possible to draw conclusions about differences in the gut-brain connectivity associations at this point, yet.

**Figure 3.**
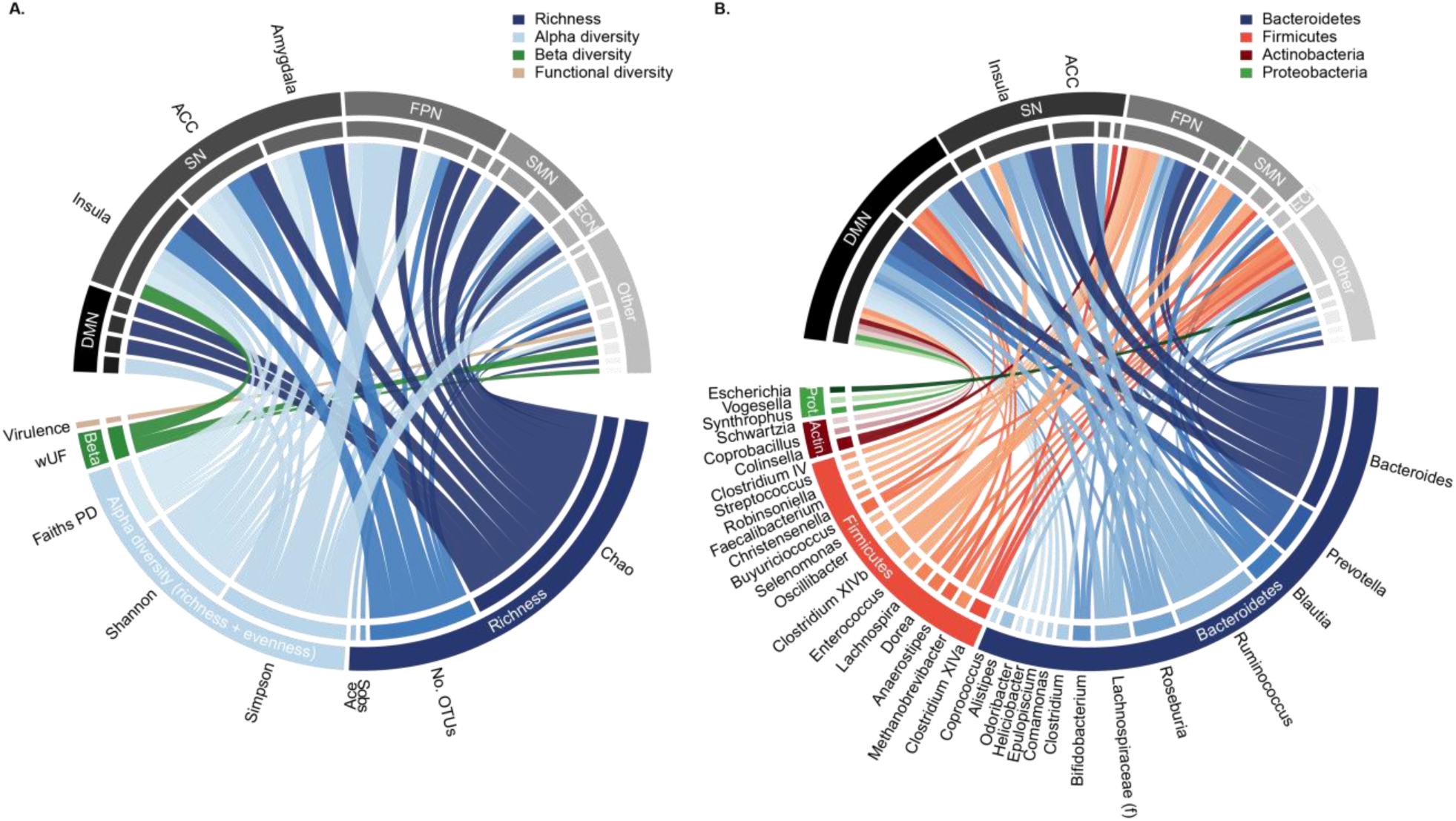
Graphic summary of the reported associations between microbial diversity and functional connectivity (**a**), and between microbial abundances and functional connectivity (**b**). Each connection in the chord diagram reflects a reported association between the abundance of that genus or diversity measure and functional connectivity. Functional connectivity is aggregated to the network level, including intra-/ and internetwork connectivity and connectivity of individual constituents of a particular network, to aid visualization. Of note, the graphical overview displays the absolute number of reported associations, skewing it towards genera and connectivity networks that are studied more frequently (a consequence of the selection bias in the reported studies). **ACC**: anterior cingulate cortex; **DMN**: default mode network; **ECN**: executive control network; **FPN**: frontoparietal network; **SMN**: sensorimotor network; SN: salience network.

Below we will first discuss the findings in relation to neurocognitive functioning and possible functional mechanisms. Finally, we will address study aspects that can explain inconsistencies in the findings, and which need to be considered to further advance the field.

### 5.2 Neurocognitive functions associated with microbiota

The current findings suggest involvement of brain networks involved in emotion-related cognition and executive functioning. The association between the gut microbiota and emotion-related functioning is a recurring topic in gut-brain research. Several of the included studies found associations between the microbiota composition and levels of anxiety, depression or negative affect, measured using questionnaires [29,30,33]. Moreover, several of the brain structures associated with the gut microbiota have a putative emotion-related function. For example, the insula, whose connectivity was associated with alpha diversity [31,34,46] and with the abundance of the genera *Roseburia* and *Bacteroides* [31, 40], is involved in socio-emotional processing, and insular brain damage can result in apathy and anxiety [47]. Moreover, the insula is involved in interoception, both emotional and visceral [8, 9]. The amygdala, whose connectivity was associated with alpha diversity and *Roseburia* abundance [30, 34], is one of the brain’s main emotion-processing structures, which is implicated in, among others, autism and anxiety [48]. Finally, the DMN, whose connectivity was associated with microbial diversity and the abundance of a wide range of bacterial genera (**Fig. 3**), is involved in mind-wandering and conceptualization of emotions [49]. Moreover, DMN connectivity is altered in depression, a disorder characterized by impairments in emotion regulation [50]. Interestingly, Kelsey and colleagues [35] reported that functional network connectivity mediated the association between microbial alpha diversity and newborn behavioral temperament, behavior that is predictive of anxiety and depression in adulthood [51]. Altogether, the associations between the gut microbiota and the brain indirectly link the gut microbiota and emotion-related behaviors through brain connectivity patterns. However, in the absence of direct, empirically tested, associations with cognition and behavior (in adults), this should be interpreted with caution.

Another set of findings suggest that the gut microbiota may be associated with executive functioning. Cai and colleagues [26] reported that frontoparietal, attention, and visual network connectivity mediated the association between higher alpha diversity and better working memory and attention, and that the orbitofrontal cortex (part of the FPN), a region involved in (emotional) decision making [52], mediated the association between *Prevotella*, *Bacteroides* and *Ruminococcus*-high clusters and response inhibition. Correspondingly, several functional networks associated with gut microbiota diversity are involved in executive functioning. For example, the frontoparietal and executive control networks (or their individual constituents) were associated with the abundance of, among others, the genera *Prevotella*, *Bacteroides* and *Blautia* [35,36,45]. These networks are mainly involved in executive control, encapsulating processes related to response inhibition, attention and working memory [6, 53].

### 5.3 Microbial function and functional pathways

Gut-to-brain signaling may occur through immunomodulation, vagal signaling or endocrine signaling [19], although there are many ways in which the gut microbiota can affect these communication pathways. Based on the taxonomic findings, one communication possibility would be through the production of short chain fatty acids (SCFAs; e.g., acetate, propionate and butyrate). For example, species within the genera *Prevotella* and *Bacteroides*, whose abundances on a genus level were associated with connectivity of a widely distributed set of brain regions and networks (**Fig. 3**), are described as main propionate and acetate-producing species [54]. Moreover, species in the genus *Blautia*, whose abundance on a genus level was associated with the connectivity of sensorimotor regions as well as network connectivity of the DMN and executive control network, are described to have propionate-producing properties [54]. Finally, species in the genus *Roseburia*, whose abundance on a genus level was associated with insular, amygdala and DMN connectivity, are among the main butyrate-producing species. Among others, SCFAs possess the ability to modulate immune activation (reviewed in [55]). In line, the association between *Roseburia* abundance and functional connectivity of the DMN and amygdala, reported by Wang and colleagues [29] and Zheng and colleagues [30], was mediated by levels of the pro-inflammatory cytokines interleukin-6 and tumor necrosis factor alpha, suggesting a role for the immune signaling pathway in the microbiota-brain connectivity communication, possibly through the production of butyrate. The involvement of this pathway may go beyond the gut, as there is preliminary evidence for a role of the immune signaling pathway in the association between functional connectivity and the oral microbiome as well [56].

Other ways through which gut-synthesized SCFAs may modulate gut-brain communication, and hereby brain connectivity, could be through maintenance of the intestinal and blood-brain barriers [55,57,58], or by traveling directly through the blood-brain barrier to the CNS [59–61], although – similar to the above-suggested role of the SCFAs - evidence directly supporting this is yet lacking.

Another way in which the gut microbiota can affect gut-brain communication is through the production of neurotransmitters and their precursors [45]. For example, species within the genera *Bacteroides* and *Bifidobacterium* and species within genera *Ruminococcus, Blautia* and *Roseburia*, whose abundances were associated with connectivity in a widespread set of brain regions and networks, are major modulators of GABA and serotonin availability in the gut [62–64]. There are several proposed pathways through which gut-derived neuroactive compounds can affect the brain. For example, there is evidence showing that both GABA and serotonin have immunomodulatory properties [65, 66], and both GABA and serotonin receptors have been located on vagal afferents to the CNS, proposing a role for the vagal signaling pathway [67, 68]. However, in the absence of direct evidence, the involvement of these pathways remains hypothetical.

Altogether, based on the taxonomic findings we can speculate about potential mechanisms. However, most taxonomic findings are reported on genus level due to the constraints of 16s rRNA sequencing. A certain genus, and even a single species within a genus, can contribute to multiple metabolite pathways. As such, our speculations should be verified through pathway analysis, preferably using higher resolution metagenomics data coupled with, when possible, bacterial culture-functional studies.

### 5.4 Recommendations for future studies

The current review identified several links between microbial genera and brain connectivity, but there is low consistency in the associations between the gut and brain. Additionally, a majority of the findings, both in microbiota and brain, were inconsistent and poorly replicated across studies. This may suggest a complex multifaceted relationship between microbial composition and brain connectivity. The limitations identified in this review underscore the need to harmonize the methodological approaches currently applied to microbiome research and (functional) brain connectivity analysis. In the following section, we will discuss important study aspects that may explain inconsistencies in the findings, and which need to be considered to further advance the field.

#### 5.4.1 Methodological comparability

The number of studies investigating the MGBA is rapidly growing, and in recent years there have been major technological advances in the field. Nevertheless, there is no golden standard as how to collect, process and analyze microbiome data. As a result, there is high inter-study variability in laboratory processing (e.g., variable region of the 16S rRNA gene and sequencing platform), pre-processing (e.g., taxonomic database, processing pipeline and prevalence filtering and data transformation) and statistical analysis approaches. The number of observed taxa and statistical outcomes can change considerably depending on the collection, storage, bioinformatic pipeline and statistical test used [69–72]. Therefore, in order to make reliable between-study comparisons it is essential to harmonize the methodological approach. To achieve this, researchers should follow standardized reporting guidelines, such as the STORMS checklist [24], on how the microbiome data was processed, follow standardized processing pipelines where possible and apply consistent and appropriate statistical approaches for analysis (e.g., accounting for the zero-inflated and compositional nature of the data).

While reducing the multiple comparison problem, additional sources of inter-study variability come from *a priori* selection of microbial genera and brain regions (in e.g., seed-based resting state analyses). Some of these genera, for example *Prevotella* and *Bacteroides*, are of common interest in studies focusing on the gut-brain communication, increasing the likelihood that the effects are observed or amplified as a result of selection bias. Moreover, *a priori* selection, both in taxonomy and brain regions, hinders meaningful comparison between study findings. At the current stage of the field, it is imperative that researchers adopt a data-driven approach to study the association between the gut microbiota and brain connectivity. Once consistent findings are obtained across studies, it is possible to use *a priori* selection of taxa and brain networks to verify hypotheses.

#### 5.4.2 Data integration and multivariate approaches

Data analysis should better acknowledge the inherent complexities of both the gut microbiota and brain connectivity data, as this will lead to a better integration of the two domains. Both the gut microbiota and the brain are complex systems, characterized by intricate relationships and interconnections [73, 74]. At the current time, most studies investigating the association between the gut microbiota and brain connectivity do so using simple bivariate association analyses, even though this is likely an oversimplification of the existing association. As a result, information concealed in the intricate relationship within and between multiple variables (i.e., interaction effects) is lost [75]. Although several studies opted for a multivariate approach for one of the two domains (i.e., differential abundance testing for the microbiome, and ICA or sparse canonical correlation analysis for brain connectivity), only one study used an integrative multivariate approach, integrating microbiota and connectivity data from microbiota and connectivity [36]. The authors identified several multivariate associations between microbial clusters and functional network connectivity.

Future studies should make an effort to integrate data from the gut microbiota and brain connectivity. Possible approaches are the Linked Independent Component Analysis (as applied in [36]) or co-inertia analysis (as applied in [37]), which allow for simultaneous factorization of data from different domains. Moreover, the systems biology approach focuses on multivariate interactions in biological systems rather than exploring each modality in parallel [76]. There are already different systems biology software programs available [77]. However, they are mainly utilized to integrate the so-called ‘omics’ techniques and, so far, there is little contribution of macroscale neurobiological measures. Nevertheless, the field of human connectomics was already proposed as an extension of systems biology to macroscale neuroscience [78–80] and could provide insight into the complex microbiota-connectivity interactions.

Another advantage to integrating more (-omics) domains would be improved mechanistic insight into the human gut-brain-axis. There have been numerous studies on possible mechanisms underlying gut-brain communication, both preclinical and in humans (reviewed in e.g., [81]). Usually, mechanisms are studied on a molecular level, without assessing how such processes would affect the brain on a larger scale. To date, only Wang and colleagues [29] and Zheng and colleagues [30] explored the mediation effect of immune activation as an underlying mechanism in the gut-brain connectivity association [29, 30]. Although we can speculate about underlying mechanisms based on taxonomic findings, it would be more informative to directly integrate mechanistic data (potentially) related with brain connectivity. For example, metagenomics can replace 16s rRNA sequencing to provide information about the functional potential of the present microbial taxa, metabolomics could provide information about bacterial metabolites that are present in the gut (e.g., SCFAs) and inflammatory markers in the blood could be investigated to provide information about immune activation.

#### 5.4.3 Association not causation

The studies discussed here were observational, from which it is not possible to infer causality nor directionality. To advance research on the human GBA, specifically for the development of gut microbiota-targeting approaches in the treatment of brain-related disorders, it is important to establish causality and directionality. Assessing directionality and causality in human trials remains a costly challenge, in which randomized controlled trials in combination with appropriate statistical models such as mediation or mendelian randomization can assist [82, 83]. For example, mediation results point towards gut-to-brain signaling, showing that immune activation (one of the MGBA signaling pathways) partially mediated the association between microbial abundance and functional brain connectivity [26, 35]. Such studies should assess if and how an intervention affects the gut microbiota or brain connectivity, which microbes are causally involved, if intervention-induced changes in the microbiota result in altered brain connectivity and – importantly – whether this is also reflected in altered cognition and behavior.

## 6 Conclusion

In this systematic review, we identified several genera as well as brain regions and networks that were repeatedly associated in microbiota-brain connectivity analyses, showing the potential of employing brain connectivity measures to gain insight in gut-brain communication. At the same time, there is still limited evidence for specificity in the microbiota-brain connectivity association. Current methodological limitations, including high inter-study variability in methodology and target population, small sample sizes, the *a priori* selection of microbial genera and brain regions of interest, and the used statistical approaches, introduce bias and thus contribute to the inconsistent findings. We know the gut-brain communication is multidimensional, and therefore a more systematic and harmonized methodology, which acknowledges this complexity, is key to further unravelling how the gut and brain communicate. To enhance comparability and replicability, future research should focus on further standardizing processing pipelines and employ data-driven multivariate analysis approaches. Moreover, interventional studies can help to clarify the causality and directionality of the reported findings.

## Data Availability

NA

## 7 Acknowledgements

The research leading to these results received funding from the European Community’s Horizon 2020 research and innovation program through the Eat2beNICE (grant agreement no. 728018), CoCA (grant agreement no. 667302) and CANDY (grant agreement no. 847818) projects.

## References

1. Sporns O, Tononi G, Kötter R. The human connectome: A structural description of the human brain. PLoS Comput Biol 2005; 1: 245–251.

2. Suárez LE, Markello RD, Betzel RF, Misic B. Linking Structure and Function in Macroscale Brain Networks. Trends Cogn Sci 2020; 24: 302–315.

3. Damoiseaux JS, Rombouts SARB, Barkhof F, Scheltens P, Stam CJ, Smith SM et al. Consistent resting-state networks across healthy subjects. Proc Natl Acad Sci 2006; 103: 13848–13853.

4. Smitha KA, Akhil Raja K, Arun KM, Rajesh PG, Thomas B, Kapilamoorthy TR et al. Resting state fMRI: A review on methods in resting state connectivity analysis and resting state networks. Neuroradiol J 2017; 30: 305.

5. Seeley WW. The Salience Network: A Neural System for Perceiving and Responding to Homeostatic Demands. J Neurosci 2019; 39: 9878–9882.

6. Marek S, Dosenbach NUF. The frontoparietal network: function, electrophysiology, and importance of individual precision mapping. Dialogues Clin Neurosci 2018; 20: 133–140.

7. Smallwood J, Bernhardt BC, Leech R, Bzdok D, Jefferies E, Margulies DS. The default mode network in cognition: a topographical perspective. Nat Rev Neurosci 2021; 22: 503–513.

8. Smith R, Alkozei A, Bao J, Smith C, Lane RD, Killgore WDS. Resting state functional connectivity correlates of emotional awareness. Neuroimage 2017; 159: 99–106.

9. Longarzo M, Quarantelli M, Aiello M, Romano M, Del Prete A, Cimminiello C et al. The influence of interoceptive awareness on functional connectivity in patients with irritable bowel syndrome. Brain Imaging Behav 2017; 11: 1117–1128.

10. Gao Y, Shuai D, Bu X, Hu X, Tang S, Zhang L et al. Impairments of large-scale functional networks in attention-deficit/hyperactivity disorder: a meta-analysis of resting-state functional connectivity. Psychol Med 2019; 49: 2475–2485.

11. Dong D, Wang Y, Chang X, Luo C, Yao D. Dysfunction of Large-Scale Brain Networks in Schizophrenia: A Meta-analysis of Resting-State Functional Connectivity. Schizophr Bull 2018; 44: 168–181.

12. Collin G, Nieto-Castanon A, Shenton ME, Pasternak O, Kelly S, Keshavan MS et al. Brain functional connectivity data enhance prediction of clinical outcome in youth at risk for psychosis. NeuroImage Clin 2020; 26: 300–311.

13. Kochunov P, Jahanshad N, Marcus D, Winkler A, Sprooten E, Nichols TE et al. Heritability of fractional anisotropy in human white matter: A comparison of Human Connectome Project and ENIGMA-DTI data. Neuroimage 2015; 111: 300–311.

14. Adhikari BM, Jahanshad N, Shukla D, Glahn DC, Blangero J, Fox PT et al. Comparison of heritability estimates on resting state fMRI connectivity phenotypes using the ENIGMA analysis pipeline. Hum Brain Mapp 2018; 39: 4893–4902.

15. Sharp PB, Sutton BP, Paul EJ, Sherepa N, Hillman CH, Cohen NJ et al. Mindfulness training induces structural connectome changes in insula networks. Sci Reports 2018 81 2018; 8: 1–10.

16. Sezer I, Pizzagalli DA, Sacchet MD. Resting-state fMRI functional connectivity and mindfulness in clinical and non-clinical contexts: A review and synthesis. Neurosci Biobehav Rev 2022; 135: 104583.

17. Jensen DEA, Leoni V, Klein-Flügge MC, Ebmeier KP, Suri S. Associations of dietary markers with brain volume and connectivity: A systematic review of MRI studies. Ageing Res Rev 2021; 70: 101360.

18. Berding K, Vlckova K, Marx W, Schellekens H, Stanton C, Clarke G et al. Diet and the Microbiota-Gut-Brain Axis: Sowing the Seeds of Good Mental Health. Adv Nutr 2021; 12: 1239–1285.

19. Cryan JF, O’riordan KJ, Cowan CSM, Sandhu K V., Bastiaanssen TFS, Boehme M et al. The Microbiota-Gut-Brain Axis. Physiol Rev 2019; 99: 1877–2013.

20. Taylor AM, Thompson S V., Edwards CG, Musaad SMA, Khan NA, Holscher HD. Associations among diet, the gastrointestinal microbiota, and negative emotional states in adults. Nutr Neurosci 2020; 23: 983–992.

21. Fattorusso A, Di Genova L, Dell’isola GB, Mencaroni E, Esposito S. Autism spectrum disorders and the gut microbiota. Nutrients 2019; 11: 521.

22. Huang TT, Lai JB, Du YL, Xu Y, Ruan LM, Hu SH. Current understanding of gut microbiota in mood disorders: An update of human studies. Front Genet 2019; 10: 1–12.

23. National Institutes of Health. Quality assessment tool for observational cohort and cross-sectional studies. 2013. [cited 2022 Sep 1]. Available from: https://www.nhlbi.nih.gov/health-topics/study-quality-assessment-tools.

24. Mirzayi C, Renson A, Furlanello C, Sansone SA, Zohra F, Elsafoury S et al. Reporting guidelines for human microbiome research: the STORMS checklist. Nat Med 2021; 27: 1885–1892.

25. Poldrack RA, Fletcher PC, Henson RN, Worsley KJ, Brett M, Nichols TE. Guidelines for reporting an fMRI study. Neuroimage 2008; 40: 409–414.

26. Cai H, Wang C, Qian Y, Zhang S, Zhang C, Zhao W et al. Large-scale functional network connectivity mediate the associations of gut microbiota with sleep quality and executive functions. Hum Brain Mapp 2021; 42: 3088–3101.

27. Zhu J, Wang C, Qian Y, Cai H, Zhang S, Zhang C et al. Multimodal neuroimaging fusion biomarkers mediate the association between gut microbiota and cognition. Prog Neuro-Psychopharmacology Biol Psychiatry 2022; 113: 110468.

28. Zhang S, Xu X, Li Q, Chen J, Liu S, Zhao W et al. Brain Network Topology and Structural– Functional Connectivity Coupling Mediate the Association Between Gut Microbiota and Cognition. Front Neurosci 2022; 16: 1–17.

29. Wang YF, Zheng LJ, Liu Y, Ye YB, Luo S, Lu GM et al. The gut microbiota-inflammation-brain axis in end-stage renal disease: Perspectives from default mode network. Theranostics 2019; 9: 8171–8181.

30. Zheng LJ, Lin L, Zhong J, Zhang Z, Ye YB, Zhang XY et al. Gut dysbiosis-influence on amygdala-based functional activity in patients with end stage renal disease: a preliminary study. Brain Imaging Behav 2020; 14: 2731–2744.

31. Curtis K, Stewart CJ, Robinson M, Molfese DL, Gosnell SN, Kosten TR et al. Insular resting state functional connectivity is associated with gut microbiota diversity. Eur J Neurosci 2019; 50: 2446–2452.

32. Hall C V., Harrison BJ, Iyer KK, Savage HS, Zakrzewski M, Simms LA, et al. Microbiota links to neural dynamics supporting threat processing. Hum Brain Mapp 2022; 43: 733–749.

33. Tillisch K, Mayer EA, Gupta A, Gill Z, Brazeilles R, Le Nevé B et al. Brain Structure and Response to Emotional Stimuli as Related to Gut Microbial Profiles in Healthy Women. Psychosom Med 2017; 79: 905–913.

34. Gao W, Salzwedel AP, Carlson AL, Xia K, Azcarate-Peril MA, Styner MA et al. Gut microbiome and brain functional connectivity in infants-a preliminary study focusing on the amygdala. Psychopharmacology (Berl) 2019; 236: 1641–1651.

35. Kelsey CM, Prescott S, McCulloch JA, Trinchieri G, Valladares TL, Dreisbach C et al. Gut microbiota composition is associated with newborn functional brain connectivity and behavioral temperament. Brain Behav Immun 2021; 91: 472–486.

36. Kohn N, Szopinska-Tokov J, Llera Arenas A, Beckmann CF, Arias-Vasquez A, Aarts E. Multivariate associative patterns between the gut microbiota and large-scale brain network connectivity. Gut Microbes 2021; 13: 2006586.

37. Li Z, Lai J, Zhang P, Ding J, Jiang J, Liu C et al. Multi-omics analyses of serum metabolome, gut microbiome and brain function reveal dysregulated microbiota-gut-brain axis in bipolar depression. Mol Psychiatry 2022;: 1–13.

38. Ahluwalia V, Betrapally NS, Hylemon PB, White MB, Gillevet PM, Unser AB et al. Impaired Gut-Liver-Brain Axis in Patients with Cirrhosis. Sci Rep 2016; 6: 1–11.

39. Jacobs JP, Gupta A, Bhatt RR, Brawer J, Gao K, Tillisch K et al. Cognitive behavioral therapy for irritable bowel syndrome induces bidirectional alterations in the brain-gut-microbiome axis associated with gastrointestinal symptom improvement. Microbiome 2021; 9: 1–14.

40. Labus JS, Osadchiy V, Hsiao EY, Tap J, Derrien M, Gupta A et al. Evidence for an association of gut microbial Clostridia with brain functional connectivity and gastrointestinal sensorimotor function in patients with irritable bowel syndrome, based on tripartite network analysis. Microbiome 2019; 7: 1–15.

41. Strandwitz P, Kim KH, Terekhova D, Liu JK, Sharma A, Levering J et al. GABA-modulating bacteria of the human gut microbiota. Nat Microbiol 2018; 4: 396–403.

42. Dong TS, Guan M, Mayer EA, Stains J, Liu C, Vora P et al. Obesity is associated with a distinct brain-gut microbiome signature that connects Prevotella and Bacteroides to the brain’s reward center. Gut Microbes 2022; 14: 1–17.

43. Dong TS, Gupta A, Jacobs JP, Lagishetty V, Gallagher E, Bhatt RR et al. Improvement in Uncontrolled Eating Behavior after Laparoscopic Sleeve Gastrectomy Is Associated with Alterations in the Brain–Gut–Microbiome Axis in Obese Women. Nutrients 2020; 12: 1– 16.

44. Hong J, Bo T, Xi L, Xu X, He N, Zhan Y et al. Reversal of Functional Brain Activity Related to Gut Microbiome and Hormones After VSG Surgery in Patients With Obesity. J Clin Endocrinol Metab 2021; 106: 3619–3633.

45. Strandwitz P. Neurotransmitter modulation by the gut microbiota. Brain Res 2018; 1693: 128–133.

46. Knight R, Vrbanac A, Taylor BC, Aksenov A, Callewaert C, Debelius J et al. Best practices for analysing microbiomes. Nat Rev Microbiol 2018; 16: 410–422.

47. Uddin LQ, Nomi JS, Hébert-Seropian B, Ghaziri J, Boucher O. Structure and function of the human insula. J Clin Neurophysiol 2017; 34: 300–306.

48. Janak PH, Tye KM. From circuits to behaviour in the amygdala. Nature 2015; 517: 284– 292.

49. Satpute AB, Lindquist KA. The Default Mode Network’s Role in Discrete Emotion. Trends Cogn Sci 2019; 23: 851–864.

50. Yan C-G, Chen X, Li L, Castellanos FX, Bai T-J, Bo Q-J et al. Reduced default mode network functional connectivity in patients with recurrent major depressive disorder. Proc Natl Acad Sci U S A 2019; 116: 9078–9083.

51. Tang A, Crawford H, Morales S, Degnan KA, Pine DS, Fox NA. Infant behavioral inhibition predicts personality and social outcomes three decades later. Proc Natl Acad Sci 2020; 117: 9800–9807.

52. Rudebeck PH, Rich EL. Current Biology Orbitofrontal cortex. Curr Biol 2018; 28: 1075– 1095.

53. Niendam TA, Laird AR, Ray KL, Dean YM, Glahn DC, Carter CS. Meta-analytic evidence for a superordinate cognitive control network subserving diverse executive functions. Cogn Affect Behav Neurosci 2012; 12: 241–268.

54. Louis P, Flint HJ. Formation of propionate and butyrate by the human colonic microbiota. Environ Microbiol 2017; 19: 29–41.

55. Venegas DP, De La Fuente MK, Landskron G, González MJ, Quera R, Dijkstra G et al. Short chain fatty acids (SCFAs)mediated gut epithelial and immune regulation and its relevance for inflammatory bowel diseases. Front Immunol 2019; 10: 277.

56. Lin D, Hutchison KE, Portillo S, Vegara V, Ellingson JM, Liu J et al. Association between the oral microbiome and brain resting state connectivity in smokers. Neuroimage 2019; 200: 121–131.

57. Silva YP, Bernardi A, Frozza RL. The Role of Short-Chain Fatty Acids From Gut Microbiota in Gut-Brain Communication. Front Endocrinol (Lausanne) 2020; 11: 1–14.

58. Hoyles L, Snelling T, Umlai UK, Nicholson JK, Carding SR, Glen RC et al. Microbiome–host systems interactions: Protective effects of propionate upon the blood–brain barrier. Microbiome 2018; 6: 1–13.

59. Frost G, Sleeth ML, Sahuri-Arisoylu M, Lizarbe B, Cerdan S, Brody L et al. The short-chain fatty acid acetate reduces appetite via a central homeostatic mechanism. Nat Commun 2014; 5: 1–11.

60. Yoo DY, Kim W, Nam SM, Kim DW, Chung JY, Choi SY et al. Synergistic effects of sodium butyrate, a histone deacetylase inhibitor, on increase of neurogenesis induced by pyridoxine and increase of neural proliferation in the mouse dentate gyrus. Neurochem Res 2011; 36: 1850–1857.

61. Erny D, De Angelis ALH, Jaitin D, Wieghofer P, Staszewski O, David E et al. Host microbiota constantly control maturation and function of microglia in the CNS. Nat Neurosci 2015; 18: 965–977.

62. Otaru N, Ye K, Mujezinovic D, Berchtold L, Constancias F, Cornejo FA et al. GABA Production by Human Intestinal Bacteroides spp.: Prevalence, Regulation, and Role in Acid Stress Tolerance. Front Microbiol 2021; 12: 1–14.

63. Barrett E, Ross RP, O’Toole PW, Fitzgerald GF, Stanton C. γ-Aminobutyric acid production by culturable bacteria from the human intestine. J Appl Microbiol 2012; 113: 411–417.

64. Yano JM, Yu K, Donaldson GP, Shastri GG, Ann P, Ma L et al. Indigenous bacteria from the gut microbiota regulate host serotonin biosynthesis. Cell 2015; 161: 264–276.

65. Bjurstöm H, Wang JY, Ericsson I, Bengtsson M, Liu Y, Kumar-Mendu S et al. GABA, a natural immunomodulator of T lymphocytes. J Neuroimmunol 2008; 205: 44–50.

66. Cloëz-Tayarani I, Changeux J-P. Nicotine and serotonin in immune regulation and inflammatory processes: a perspective. J Leukoc Biol 2007; 81: 599–606.

67. Breit S, Kupferberg A, Rogler G, Hasler G. Vagus nerve as modulator of the brain-gut axis in psychiatric and inflammatory disorders. Front Psychiatry 2018; 9: 1–15.

68. Cawthon CR, de La Serre CB. Gut bacteria interaction with vagal afferents. Brain Res 2018; 1693: 134–139.

69. Boers SA, Jansen R, Hays JP. Understanding and overcoming the pitfalls and biases of next-generation sequencing (NGS) methods for use in the routine clinical microbiological diagnostic laboratory. Eur J Clin Microbiol Infect Dis 2019; 38: 1059–1070.

70. Nearing JT, Douglas GM, Hayes MG, MacDonald J, Desai DK, Allward N et al. Microbiome differential abundance methods produce different results across 38 datasets. Nat Commun 2022; 13: 1–16.

71. Teng F, Darveekaran Nair SS, Zhu P, Li S, Huang S, Li X et al. Impact of DNA extraction method and targeted 16S-rRNA hypervariable region on oral microbiota profiling. Sci Rep 2018; 8: 1–12.

72. Szopinska-Tokov J, Bloemendaal M, Boekhorst J, Hermes G DA, Ederveen T, Vlaming P et al. A comparison of bioinformatics pipelines for compositional analysis of the human gut microbiome (submitted).

73. Thursby E, Juge N. Introduction to the human gut microbiota. Biochem J 2017; 474: 1823–1836.

74. Kelly C, Biswal BB, Craddock RC, Castellanos FX, Milham MP. Characterizing variation in the functional connectome: promise and pitfalls. Trends Cogn Sci 2012; 16: 181–188.

75. Lahat D, Adali T, Jutten C. Multimodal Data Fusion: An Overview of Methods, Challenges, and Prospects. Proc IEEE 2015; 103: 1449–1477.

76. Tavassoly I, Goldfarb J, Iyengar R. Systems biology primer: The basic methods and approaches. Essays Biochem 2018; 62: 487–500.

77. De Souza HSP, Fiocchi C, Iliopoulos D. The IBD interactome: An integrated view of aetiology, pathogenesis and therapy. Nat Rev Gastroenterol Hepatol 2017; 14: 739–749.

78. Bigler ED. Systems biology, neuroimaging, neuropsychology, neuroconnectivity and traumatic brain injury. Front Syst Neurosci 2016; 10: 1–23.

79. Holzinger A, Haibe-Kains B, Jurisica I. Why imaging data alone is not enough: AI-based integration of imaging, omics, and clinical data. Eur J Nucl Med Mol Imaging 2019; 46: 2722–2730.

80. Sporns O. The human connectome: Origins and challenges. Neuroimage 2013; 80: 53–61.

81. Martin CR, Osadchiy V, Kalani A, Mayer EA. The Brain-Gut-Microbiome Axis. Cell Mol Gastroenterol Hepatol 2018; 6: 133–148.

82. Lv BM, Quan Y, Zhang HY. Causal Inference in Microbiome Medicine: Principles and Applications. Trends Microbiol 2021; 29: 736–746.

83. Chaudhari SN, McCurry MD, Devlin AS. Chains of evidence from correlations to causal molecules in microbiome-linked diseases. Nat Chem Biol 2021; 17: 1046–1056.

84. Wang WL, Xu SY, Ren ZG, Tao L, Jiang JW, Zheng S Sen. Application of metagenomics in the human gut microbiome. World J Gastroenterol 2015; 21: 803.

85. Hugerth LW, Andersson AF. Analysing microbial community composition through amplicon sequencing: From sampling to hypothesis testing. Front Microbiol 2017; 8: 1561.

86. Emsell L, Van Hecke W, Tournier JD. Introduction to diffusion tensor imaging. In: Diffusion Tensor Imaging: A Practical Handbook. Springer: New York, 2016, pp 7–19.

87. Bijsterbosch J, Smith SM, Beckmann CF. Introduction to Resting State fMRI Functional Connectivity. Oxford Neuroimaging Prim 2017.

88. Beckmann CF, DeLuca M, Devlin JT, Smith SM. Investigations into resting-state connectivity using independent component analysis. Philos Trans R Soc B Biol Sci 2005; 360: 1001–1013.

89. Hevey D. Network analysis: a brief overview and tutorial. Heal Psychol Behav Med 2018; 6: 301–328.

