## Supplemental Table 1 for "A systematic review exploring the association between the human gut microbiota and brain connectivity in health and disease"

**Overview over the reported findings per study for microbial diversity, composition and clusters**

1: Association reported
0: No association found
No association assessed

| Category | Order | Family | Measure / Genus | Curtis (2019) | Cai (2021) / Zhu (2022) / Zhang (2022) | Hall (2021) | Kohn (2021) | Tillisch (2017) | Gao (2019) | Kelsey (2021) | Wang (2019) / Zheng (2020) | Li (2022) | Dong (2022) | Dong (2020) | Strandwitz (2019) | Ahluwalia (2016) | Labus (2019) | Hong (2021) | Jacobs (2021) | Studies reported | Studies assessed | % |
|---|---|---|---|---|---|---|---|---|---|---|---|---|---|---|---|---|---|---|---|---|---|---|
| Microbial diversity | **Richness** | | | | | | | | | | | | | | | | | | | | | |
| | | | Number of OTUs | 1 | | | | | 1 | | | | | | | | | | | 2 | 2 | 100% |
| | | | Sobs | | 1 | | | | | | | | | | | | | | | 1 | 1 | 100% |
| | | | Ace | | 1 | | | | | | | | | | | | | | | 1 | 1 | 100% |
| | | | Chao | | 1 | | | | 1 | 1 | | | | | | | | | | 3 | 3 | 100% |
| | **Alpha Diversity** | | | | | | | | | | | | | | | | | | | | | |
| | | | Shannon | 0 | 1 | | | | 1 | 1 | | | | 0 | | | | | | 3 | 5 | 60% |
| | | | Simpson | | 1 | 1 | | | | | | | | | | | | | | 2 | 2 | 100% |
| | | | Faiths PD | | | | | | 1 | | | | | | | | | | | 1 | 1 | 100% |
| | **Beta Diversity** | | | | | | | | | | | | | | | | | | | | | |
| | | | Weighted UniFrac | 1 | | | | | | | | | | | | | | | | 1 | 1 | 100% |
| Microbial composition | **Order** | **Family** | **Genus** | | | | | | | | | | | | | | | | | | | |
| | Acidaminicoccales | Acidaminococcaceae | Phascolarctobacterium | | | | 0 | | | 0 | | 1 | | 0 | | | | | | 1 | 4 | 25% |
| | Actinomycetales | Actinomycetaceae | Actinomyces | | | | 0 | | | 0 | 0 | 0 | 1 | 0 | | | | | | 1 | 6 | 17% |
| | Bacteroidales | Prevotellaceae | Alistipes | | | | 0 | | | 0 | | 1 | | 1 | | | | | | 2 | 4 | 50% |
| | Bacteroidales | Bacteroidaceae | Bacteroides | 1 | | 1 | 1 | 0 | | 1 | | 1 | 1 | 1 | 1 | | | 0 | 1 | 9 | 11 | 82% |
| | Bacteroidales | Porphyromonadaceae | Methanobrevibacter | | | | 0 | | | 0 | | 0 | | 1 | | | | | | 1 | 4 | 25% |
| | Bacteroidales | Odoribacteraceae | Odoribacter | | | | 0 | | | 0 | 1 | 0 | 0 | 0 | | | | | | 1 | 6 | 17% |
| | Bacteroidales | Tannerellaceae | Parabacteroides | | | | 0 | | | 0 | 0 | 1 | 0 | 0 | | | | | | 1 | 6 | 17% |
| | Bacteroidales | Prevotellaceae | Prevotella | 1 | | 0 | 1 | 1 | | 1 | 1 | 1 | 0 | 0 | | | | | 0 | 6 | 10 | 60% |
| | Bifidobacteriales | Bifidobacteriaceae | Bifidobacterium | | | | 1 | | | 1 | | | | 0 | | | | | | 2 | 3 | 67% |
| | Burkholderiales | Comamonadaceae | Comamonas | | | | 0 | | | 0 | 1 | | | 0 | | | | | | 1 | 4 | 25% |
| | Burkholderiales | Sutterellaceae | Sutterella | | | | 0 | | | 0 | | 1 | | 0 | | | | | | 1 | 4 | 25% |
| | Campylobacterales | Helicobacteraceae | Heliciobacter | | | | 0 | | | 0 | 1 | 1 | | 0 | | | | | | 2 | 5 | 40% |
| | Christensenellales | Christensenellaceae | Christensenella | | | | 1 | | | 0 | | | | 0 | | | | | | 1 | 3 | 33% |
| | Clostridiales | Lachnospiraceae | Anaerostipes | | | | 0 | | | 0 | | | | 1 | | | 0 | | | 1 | 4 | 25% |
| | Clostridiales | Lachnospiraceae | Blautia | | | | 1 | | | 0 | | 1 | | 0 | | | 1 | | | 3 | 5 | 60% |
| | Clostridiales | Clostridiaceae | Butyricicoccus | | | | 0 | | | 0 | | | | 1 | | | | | | 1 | 3 | 33% |
| | Clostridiales | Catabacteriaceae | Catabacter | | | | 0 | | | 0 | | 1 | | 0 | | | | | | 1 | 4 | 25% |
| | Clostridiales | Ruminococcaceae | Clostridium | | | | 0 | | | 1 | 0 | 1 | | 0 | | | 1 | 0 | | 3 | 7 | 43% |
| | Clostridiales | Lachnospiraceae | Coprococcus | | | | 0 | | | 0 | 0 | | | 0 | | | 1 | | | 1 | 5 | 20% |
| | Clostridiales | Veillonellaceae | Dorea | | | | 0 | | | 0 | | | | 1 | | | | | | 1 | 3 | 33% |
| | Clostridiales | Eubacteriaceae | Eubacterium | | | | 0 | | | 0 | | 1 | 1 | 0 | | | | | | 2 | 5 | 40% |
| | Clostridiales | | Epulopiscium | | | | 0 | | | 0 | 1 | | | 0 | | | | | | 1 | 4 | 25% |
| | Clostridiales | Ruminococcaceae | Faecalibacterium | | | | 1 | | | 0 | | 1 | | 0 | | | 0 | | | 2 | 5 | 40% |
| | Clostridiales | Lachnospiraceae | Lachnospira | | | | 0 | | | 0 | 0 | 0 | | 1 | | | 0 | | | 1 | 6 | 17% |
| | Clostridiales | Ruminococcaceae | Oscillibacter | | | | 0 | | | 0 | | 1 | | 0 | | | 1 | | | 2 | 5 | 40% |
| | Clostridiales | Lachnospiraceae | Robinsoniella | | | | 0 | | | 1 | | 1 | | 0 | | | 0 | | | 2 | 5 | 40% |
| | Clostridiales | Lachnospiraceae | Roseburia | | | | 0 | | | 0 | 1 | 1 | | 0 | | | 1 | | 0 | 3 | 7 | 43% |
| | Clostridiales | Ruminococcaceae | Ruminococcus | | | 1 | 1 | | | 0 | 0 | 1 | | 1 | | | 0 | | | 4 | 7 | 57% |
| | Coriobacteriales | Coriobacteriaceae | Collinsella | | | | 0 | | | 1 | 1 | 1 | | 0 | | | | | | 3 | 5 | 60% |
| | Enterobacterales | Enterobacteriaceae | Enterobacter | | | | 0 | | | 0 | | 1 | | 0 | | | | | | 1 | 4 | 25% |
| | Enterobacterales | Enterobacteriaceae | Escherichia | | | | 0 | | | 1 | | | | 0 | | | | | | 1 | 3 | 33% |
| | Erysipelotrichales | Erysipelotrichaceae | Coprobacillus | | | | 0 | | | 0 | 1 | 1 | | 0 | | | | | | 2 | 5 | 40% |
| | Eubacteriales | Lachnospiraceae | Lachnoclostridium | | | | 0 | | | 0 | | 1 | | 0 | | | | | | 1 | 4 | 25% |
| | Eubacteriales | Lachnospiraceae | Oribacterium | | | | 0 | | | 0 | 0 | | 1 | 0 | | | | | | 1 | 5 | 20% |
| | Fusobacteriales | Fusobacteriaceae | Fusobacterium | | | | 0 | | | 0 | | 1 | 1 | 0 | | | | | | 2 | 5 | 40% |
| | Lactobacillales | Enterococcaceae | Enterococcus | | | | 0 | | | 1 | | 1 | | 0 | | | | | | 2 | 4 | 50% |
| | Lactobacillales | Streptococcaceae | Streptococcus | | | | 0 | | | 1 | | 1 | 0 | 0 | | | | 0 | | 2 | 6 | 33% |
| | Neisseriales | Neisseriaceae | Vogesella | | | | 0 | | | 0 | 1 | | | 0 | | | | | | 1 | 4 | 25% |
| | Selenomonadales | Selenomonadaceae | Schwartzia | | | | 0 | | | 0 | 1 | | | 0 | | | | | | 1 | 4 | 25% |
| | Selenomonadales | Selenomonadaceae | Selenomonas | | | | 0 | | | 0 | 1 | | | 0 | | | | | | 1 | 4 | 25% |
| | Syntrophobacterales | Syntrophaceae | Synthrophus | | | | 0 | | | 0 | 1 | | | 0 | | | | | | 1 | 4 | 25% |
| Clusters | | | Bacteroides cluster | | 1 | | | 1 | | | 1 | | | | | | | | | 3 | 3 | 100% |
| | | | Prevotella cluster | | 1 | | | 1 | | | 1 | | | | | | | | | 3 | 3 | 100% |
| | | | Ruminococcus cluster | | 1 | | | | | | | | | | | | | | | 1 | 1 | 100% |
| Ratio | | | Prevotella/Bacteroides | | | | | | | | | | 1 | | | | | | | 1 | 1 | 100% |
