## Supplemental Table 2 for "A systematic review exploring the association between the human gut microbiota and brain connectivity in health and disease"

Overview of the reported findings per study for functional and structural connectivity, aggregated to the network level

1: Association reported
0: No association found
No association assessed

| | Region | Curtis (2019) | Cai (2021) / Zhu (2022) / Zhang (2022) | Hall (2021) | Kohn (2021) | Tillisch (2017) | Gao (2019) | Kelsey (2021) | Wang (2019) / Zheng (2020) | Li (2022) | Dong (2022) | Dong (2020) | Strandwitz (2019) | Ahluwalia (2016) | Labus (2019) | Hong (2021) | Jacobs (2021) | Studies reported | Studies assessed | % |
|---|---|---|---|---|---|---|---|---|---|---|---|---|---|---|---|---|---|---|---|---|
| Functional connectivity | **Auditory network** | | 1 | | | | | | | | | | | | | | | 1 | 1 | 100% |
| | **Default mode network** | | 1 | | 1 | | | 1 | 1 | 1 | | | 1 | | | | | 6 | 6 | 100% |
| | Angular gyrus | 0 | 1 | | | | 0 | | 0 | 1 | | | | | | | | 2 | 5 | 40% |
| | Medial frontal cortex | 0 | 0 | | | | 0 | | 1 | 1 | | | | | | | | 2 | 5 | 40% |
| | Precuneus | 0 | 1 | | | | 0 | | 0 | 1 | | 1 | | | | | | 3 | 6 | 50% |
| | Ventromedial prefrontal cortex | 0 | 1 | | | | 0 | | 0 | | | | | | | | | 1 | 4 | 25% |
| | **Dorsal attention network** | | 1 | | | | | | | | | | | | | | | 1 | 1 | 100% |
| | **Executive control network** | | 1 | | 1 | | | | | | | | | | | | | 2 | 2 | 100% |
| | **Frontoparietal network** | | 1 | | 1 | | | 1 | | 1 | | | | | | | | 4 | 4 | 100% |
| | Dorsolateral prefrontal cortex | 0 | 1 | | | | 0 | | 0 | 1 | | | 1 | | | | | 3 | 6 | 50% |
| | Inferior parietal lobe | 0 | 0 | | | | 1 | | 1 | | | | | | | | | 2 | 4 | 50% |
| | Medial cingulate cortex | 0 | 1 | | | | 0 | | 0 | | | | | | | | 0 | 1 | 5 | 20% |
| | **Homologous-interhemispheric network** | | | | | | | 1 | | | | | | | | | | 1 | 1 | 100% |
| | **Salience network** | | 0 | | 1 | | | | | 1 | | | | | | | | 2 | 3 | 67% |
| | Amygdala | 0 | 0 | | | | 1 | | 1 | 1 | | | | | | | 0 | 3 | 6 | 50% |
| | Anterior cingulate cortex | 0 | 1 | 1 | 1 | | 1 | | 1 | 1 | | | | | | | 0 | 6 | 8 | 75% |
| | Insula | 1 | 1 | 1 | | | 1 | | 0 | 1 | | | | | 1 | | 0 | 6 | 8 | 75% |
| | Nucleus accumbens | 0 | 0 | | | | 0 | | 0 | 1 | 1 | | | | 1 | | | 3 | 7 | 43% |
| | **Sensorimotor network** | 0 | 1 | | | | 0 | | 0 | 1 | | | | | | | | 2 | 5 | 40% |
| | M1 | 0 | 1 | | | | 0 | | 0 | | | | | | 1 | | | 2 | 5 | 40% |
| | S1 | 0 | 1 | | | | 0 | | 0 | | | | | | 1 | | | 2 | 5 | 40% |
| | S2 | 0 | 0 | | | | 1 | | 0 | | | | | | 1 | | | 2 | 5 | 40% |
| | Supplementary motor area | 0 | 0 | | | | 0 | | 0 | 1 | | | | | | 0 | | 1 | 6 | 17% |
| | **Visual network** | | 1 | | | | | | | 1 | | | | | | | | 2 | 2 | 100% |
| | **Other** | | | | | | | | | | | | | | | | | | | |
| | Brainstem | 0 | 0 | | | | 0 | | 0 | | 1 | | | | | | 1 | 2 | 6 | 33% |
| | Caudate | 0 | 0 | | | | 0 | | 0 | 1 | | | | | 1 | | | 2 | 6 | 33% |
| | Cerebellum | 1 | 0 | | | | 0 | | 0 | | | | | | | | | 1 | 4 | 25% |
| | Dorsomedial prefrontal cortex | 0 | 1 | | | | 0 | | 0 | | | | | | | | | 1 | 4 | 25% |
| | Frontal pole | 1 | 0 | | | | 0 | | 0 | 1 | | | | | | | | 2 | 5 | 40% |
| | Fusiform gyrus | 0 | 1 | | | | 0 | | 0 | | | | | | | | | 1 | 4 | 25% |
| | Herschl's gyrus | 0 | 1 | | | | 0 | | 0 | | | | | | | | 0 | 1 | 5 | 20% |
| | Hippocampus | 0 | 0 | | | | 0 | | 0 | 1 | | | | | | | | 1 | 5 | 20% |
| | Lateral occipital cortex | 0 | 1 | | | | 0 | | 0 | 1 | | | | | | | | 2 | 5 | 40% |
| | Lateral prefrontal cortex | 0 | 1 | | | | 0 | | 0 | 1 | | | | | | | | 2 | 5 | 40% |
| | Lateral temporal cortex | 0 | 1 | | | | 0 | | 0 | | | | | | | | | 1 | 4 | 25% |
| | Lingual gyrus | 1 | 0 | | | | 0 | | 0 | 1 | | | | | | | | 2 | 5 | 40% |
| | Occipital cortex | 1 | 0 | | | | 0 | | 0 | 1 | | | | | | | | 2 | 5 | 40% |
| | Operculum | 1 | 0 | | | | 0 | | 0 | 1 | | | | | | | | 2 | 5 | 40% |
| | Pallidum | 0 | 0 | | | | 0 | | 0 | 1 | | | | | 1 | | | 2 | 6 | 33% |
| | Putamen | 0 | 0 | | | | 0 | | 0 | 1 | | 1 | | | 0 | 0 | | 2 | 8 | 25% |
| | Superior parietal lobule | 0 | 1 | | | | 0 | | 0 | 1 | | | | | | | | 2 | 5 | 40% |
| | Superior temporal gyrus | 0 | 1 | | | | 0 | | 0 | 1 | | | | | | | 1 | 3 | 6 | 50% |
| | Temporal pole | 0 | 1 | | | | 0 | | 0 | 1 | | | | | | | | 2 | 5 | 40% |
| | Thalamus | 0 | 0 | | | | 1 | | 0 | 1 | | | | | 0 | | | 2 | 6 | 33% |
| Structural connectivity | Corpus callosum | | 1 | | | | | | | | | | | 1 | | | | 2 | 2 | 100% |
| | Internal capsule | | 1 | | | | | | | | | | | 1 | | | | 2 | 2 | 100% |
| | Optic radiation | | 1 | | | | | | | | | | | | | | | 1 | 1 | 100% |
| | Temportal juxtacortical white matter | | 1 | | | | | | | | | | | | | | | 1 | 1 | 100% |
| | Occipital juxtacortical white matter | | 1 | | | | | | | | | | | | | | | 1 | 1 | 100% |
| | Frontal white matter | | 1 | | | | | | | | | | | 1 | | | | 2 | 2 | 100% |
| | Parietal white matter | | 1 | | | | | | | | | | | | | | | 1 | 1 | 100% |
| | Middle frontal gyrus-central sulcus | | | | | 1 | | | | | | | | | | | | 1 | 1 | 100% |
| | Amygdala-caudate | | | | | 1 | | | | | | | | | | | | 1 | 1 | 100% |
| | ACC-pallidum | | | | | 1 | | | | | | | | | | | | 1 | 1 | 100% |
| | Fusiform gyrus-inferior temporal gyrus | | | | | 1 | | | | | | | | | | | | 1 | 1 | 100% |
| | Anterior transverse collateral sulcus-inferior temporal sulcus | | | | | 1 | | | | | | | | | | | | 1 | 1 | 100% |
| | Inferior temporal sulcus-parallel sulcus | | | | | 1 | | | | | | | | | | | | 1 | 1 | 100% |
| | Thalamus-pericallosal sulcus | | | | | 1 | | | | | | | | | | | | 1 | 1 | 100% |
| | Posterior ramus of the lateral sulcus-temporal pole | | | | | 1 | | | | | | | | | | | | 1 | 1 | 100% |
| | Thamalus-temporal pole | | | | | 1 | | | | | | | | | | | | 1 | 1 | 100% |
| | Posterior mid cingulate gyrus/sulcus-central sulcus | | | | | 1 | | | | | | | | | | | | 1 | 1 | 100% |
| | Inferior longitudinal fasciculi | | 0 | | | | | | | | | | | 1 | | | | 1 | 2 | 50% |
| | Inferior temporal gyrus | | 1 | | | | | | | | | | | | | | | 1 | 1 | 100% |
| | Insula | | 0 | | | | | | | | | | | | | | | 0 | 1 | 0% |
| | Caudate nucleus | | 1 | | | | | | | | | | | | | | | 1 | 1 | 100% |
| | Corticospinal tracts | | 0 | | | | | | | | | | | 0 | | | | 0 | 2 | 0% |
| | Posterior white matter | | 0 | | | | | | | | | | | 1 | | | | 1 | 2 | 50% |
| | Posterior cingulare gyrus | | 1 | | | | | | | | | | | | | | | 1 | 1 | 100% |
| | Superior longitudinal fasciculi | | 0 | | | | | | | | | | | 0 | | | | 0 | 2 | 0% |
| | Supramarginal gyrus | | 1 | | | | | | | | | | | | | | | 1 | 1 | 100% |
| | Uncinate fasciculi | | 0 | | | | | | | | | | | 1 | | | | 1 | 2 | 50% |
| SC - FC coupling | Anterior cingulate cortex | | 1 | | | | | | | | | | | | | | | 1 | 1 | 100% |
| | Hippocampus | | 1 | | | | | | | | | | | | | | | 1 | 1 | 100% |
| | Fusiform gyrus | | 1 | | | | | | | | | | | | | | | 1 | 1 | 100% |
| | Medial superior frontal gyrus | | 1 | | | | | | | | | | | | | | | 1 | 1 | 100% |
| | Supramarginal gyrus | | 1 | | | | | | | | | | | | | | | 1 | 1 | 100% |
