## Supplemental Table 3 for "A systematic review exploring the association between the human gut microbiota and brain connectivity in health and disease"

**Study Quality Assessment Tools for Observational Cohort and Cross-Sectional studies by National Heart, Lung and Blood Institute (2013), adapted to include criteria for case-control and before-after studies with no control group. Adaptations made by the authors are specified in the column "guidance"**

| | Question | Guidance |
|---|---|---|
| **Q1** | Was the research question or objective in this paper clearly stated? | Did the authors describe their goal in conducting this research? Is it easy to understand what they were looking to find? This issue is important for any scientific paper of any type. Higher quality scientific research explicitly defines a research question. |
| **Q2** | Was the study population clearly specified and defined?<br><br>*For case-control studies: Were the cases clearly defined and differentiated from controls?* | Did the authors describe the group of people from which the study participants were selected or recruited, using demographics, location, and time period? If you were to conduct this study again, would you know who to recruit, from where, and from what time period? Is the cohort population free of the outcomes of interest at the time they were recruited?<br><br>*For case-control studies: was a specific description of "case" and "control" provided? Is there a discussion of the validity of the case and control definitions and the processes or tools used to identify study participants as such? Were tools or methods accurate, reliable, and objective?* |
| **Q3** | Was the participation rate of eligible persons at least 50%?<br><br>*For case-control studies: if less than 100 percent of eligible cases and/or controls were selected for the study, were the cases and/or controls randomly selected from those eligible?* | If fewer than 50% of eligible persons participated in the study, then there is concern that the study population does not adequately represent the target population. This increases the risk of bias.<br><br>*For case-control studies: if a case-control study did not use 100 percent of eligible cases and/or controls, did the authors indicate that random sampling was used to select controls? If investigators included all eligible cases and controls as study participants, this criterion is marked "NA"* |
| **Q4** | Were all the subjects selected or recruited from the same or similar populations (including the same time period)? Were inclusion and exclusion criteria for being in the study prespecified and applied uniformly to all participants?<br><br>*For case-control studies: Were controls selected or recruited from the same or similar population that gave rise to the cases (including the same timeframe)?* | Were the inclusion and exclusion criteria developed prior to recruitment or selection of the study population? Were the same underlying criteria used for all of the subjects involved?<br><br>To determine whether cases and controls were recruited from the same population, one can ask hypothetically, "If a control was to develop the outcome of interest, would that person have been eligible to become a case?" |
| **Q5** | Was there use of concurrent controls? | A concurrent control is a control selected at the time another person became a case, usually on the same day. This means that one or more controls are recruited or selected from the population without the outcome of interest at the time a case is diagnosed.<br><br><u>Authors note</u>*: this criterium was considered "NA" for all non-case-control studies* |
| **Q6** | Was a sample size justification, power description, or variance and effect estimates provided? | Did the authors present their reasons for selecting or recruiting the number of people included or analyzed? Do they note or discuss the statistical power of the study? This question is about whether or not the study had enough participants to detect an association if one truly existed. A paragraph in the methods section of the article may explain the sample size needed to detect a hypothesized difference in outcomes. You may also find a discussion of power in the discussion section. Sometimes estimates of variance and/or estimates of effect size are given, instead of sample size calculations. In any of these cases, the answer would be "yes."<br><br>However, observational (cohort) studies often do not report anything about power or sample sizes because the analyses are exploratory in nature. In this case, the answer would be "no." This is not a "fatal flaw." It just may indicate that attention was not paid to whether the study was sufficiently sized to answer a prespecified question–i.e., it may have been an exploratory, hypothesis-generating study. |
| **Q7** | For the analyses in this paper, were the exposure(s) of interest measured prior to the outcome(s) being measured? | This question is important because, in order to determine whether an exposure causes an outcome, the exposure must come before the outcome. Sometimes cross-sectional studies are conducted (or cross-sectional analyses of cohort-study data), where the exposures and outcomes are measured during the same timeframe. As a result, cross-sectional analyses provide weaker evidence than regular cohort studies regarding a potential causal relationship between exposures and outcomes.<br><br><u>Authors note</u>*: this criterium was marked "no" for all analyses based on one timepoint of measurement* |
| **Q8** | Was the timeframe sufficient so that one could reasonably expect to see an association between exposure and outcome if it existed? | Cross-sectional analyses allow no time to see an effect, since the exposures and outcomes are assessed at the same time.<br><br><u>Authors note</u>*: this criterium was marked "no" for all analyses based on one timepoint of measurement* |
| **Q9** | For exposures that can vary in amount or level, did the study examine different levels of the exposure as related to the outcome (e.g., categories of exposure, or exposure measured as continuous variable)? | If the exposure can be defined as a range, were multiple categories of that exposure assessed? Sometimes discrete categories of exposure are not used, but instead exposures are measured as continuous variables |

| | Question | Guidance |
|---|---|---|
| **Q10** | Were the exposure measures (independent variables) clearly defined, valid, reliable, and implemented consistently across all study participants? | Were the exposure measures defined in detail? Were the tools or methods used to measure exposure accurate and reliable–for example, have they been validated or are they objective? This issue is important as it influences confidence in the reported exposures. When exposures are measured with less accuracy or validity, it is harder to see an association between exposure and outcome even if one exists.<br><br>*Author's note: in this review, we considered the gut microbiota composition to be the exposure. This criterium was marked using the method section of the STORMS checklist (v1.03)* |
| **Q11** | Was the exposure(s) assessed more than once over time?<br>*For intervention studies: were outcome measures of interest taken multiple times before the intervention and multiple times after the intervention (i.e., did they use an interrupted time-series design)?* | Was the exposure for each person measured more than once during the course of the study period? Multiple measurements with the same result increase our confidence that the exposure status was correctly classified. Also, multiple measurements enable investigators to look at changes in exposure over time. |
| **Q12** | Were the outcome measures (dependent variables) clearly defined, valid, reliable, and implemented consistently across all study participants? | Were the outcomes defined in detail? Were the tools or methods for measuring outcomes accurate and reliable–for example, have they been validated or are they objective? This issue is important because it influences confidence in the validity of study results.<br><br>*Author's note: in this review, we considered the structural or functional connectivity to be the outcome. This criterium was marked using an adaptation of the guidelines/checklist by Poldrack et al (2008)* |
| **Q13** | Were the outcome assessors blinded to the exposure status of participants?<br>*For case-control studies: Were the assessors of exposure/risk blinded to the case or control status of participants?* | Blinding means that outcome assessors did not know whether the participant was exposed or unexposed. It is also sometimes called "masking." If blinding was not possible, which is sometimes the case, mark "NA" and explain the potential for bias.<br><br>*Author's note: in this review the exposure variable is the gut microbiota composition. For all non-case-control/non-intervention studies, this criterion was marked "NA" as blinding for the gut microbiota composition is not of added value* |
| **Q14** | Was loss to follow-up after baseline 20% or less? | Higher overall followup rates are always better than lower followup rates, even though higher rates are expected in shorter studies, whereas lower overall followup rates are often seen in studies of longer duration. Usually, an acceptable overall followup rate is considered 80 percent or more of participants whose exposures were measured at baseline<br><br>*Author's note: this criterion was marked "NA" for all studies with only one timepoint of measurement* |
| **Q15** | Were key potential confounding variables measured and adjusted statistically for their impact on the relationship between exposure(s) and outcome(s)? | Were key potential confounding variables measured and adjusted for, such as by statistical adjustment for baseline differences? Logistic regression or other regression methods are often used to account for the influence of variables not of interest. All key factors that may be associated both with the exposure of interest and the outcome–that are not of interest to the research question–should be controlled for in the analyses.<br><br>*Author's note: in this review, age and sex were considered key confounding variables.* |
| **Q16** | Were the association analyses between exposure and outcome clearly defined, valid, reliable and properly implemented?<br><br>*For interventional studies: did the statistical methods examine changes in outcome measures from before to after the intervention? Were statistical tests done that provided p values for the pre-to-post changes?* | *Author's note: this review focusses on the association between the exposure (i.e. the gut microbiota composition) and the outcome (i.e. functional and/or structural connectivity). Hence, this criterion was added to assess the validity and reliability of the used analytical approach. Was the statistical approach defined in detail? Were the tools or methods for testing the association between exposure and outcome accurate and reliable–for example, have they been validated or are they objective? Was a correction for multiple testing implemented?*<br><br>*For interventional studies: were formal statistical tests used to assess the significance of the changes in the outcome measures between the before and after time periods? The reported study results should present values for statistical tests, such as p values, to document the statistical significance (or lack thereof) for the changes in the outcome measures found in the study.* |
