## Supplemental Table 4 for "A systematic review exploring the association between the human gut microbiota and brain connectivity in health and disease"

**The quality assessment of the microbiome measurement per study, using the STORMS (v1.03) criteria**

| Number | Item | Recommendation | Additional Guidance | Curtis (2019) | Cai (2021) / Zhu (2022) / Zhang (2022) | Hall (2021) | Kohn (2021) | Tillisch (2017) | Gao (2019) | Kelsey (2021) | Wang (2019) / Zheng (2020) | Dong (2022) | Dong (2020) | Strandwitz (2019) | Ahluwalia (2016) | Labus (2019) | Li (2022) | Hong (2021) | Jacobs (2021) |
|---|---|---|---|---|---|---|---|---|---|---|---|---|---|---|---|---|---|---|---|
| 4,0 | Laboratory methods | State the laboratory/center where laboratory work was done. | Provide a reference to complete lab protocols if previously published elsewhere such as on protocols.io. Note any modifications of lab protocols and the reason for protocol modifications. | no | no | yes | yes | yes | no | yes | no | no | no | yes | no | yes | no | yes | yes |
| 4,1 | Specimen collection | State the body site(s) sampled from and how specimens were collected. | Use terms from the Uber-anatomy Ontology (https://www.ebi.ac.uk/ols/ontologies/uberon) to describe body sites in a standardized format. | yes | yes | yes | yes | yes | yes | yes | yes | no | no | yes | no | yes | yes | yes | yes |
| 4,2 | Shipping | Describe how samples were stored and shipped to the laboratory. | Include length of time from collection to receipt by the lab and if temperature control was used during shipping. | no | yes | yes | yes | yes | yes | yes | no | no | no | yes | no | yes | yes | yes | yes |
| 4,3 | Storage | Describe how the laboratory stored samples, including time between collection and storage and any preservation buffers or refrigeration used. | State where each procedure or lot of samples was done if not all in the same place.<br><br>Include reagent/lot/catalogue #s for storage buffers. | no | yes | yes | yes | yes | yes | yes | yes | yes | no | yes | no | no | yes | yes | yes |
| 4,4 | DNA extraction | Provide DNA extraction method, including kit and version if relevant. | If any DNA quantification methods were used prior to DNA amplification or at the pooling step of library preparation, state so here. | yes | yes | yes | yes | yes | yes | yes | yes | yes | no | yes | yes | yes | yes | yes | yes |
| 4,5 | Human DNA sequence depletion or microbial DNA enrichment | Describe whether human DNA sequence depletion or enrichment of microbial or viral DNA was performed. | | yes | yes | yes | yes | yes | yes | yes | yes | yes | yes | yes | yes | yes | yes | yes | yes |
| 4,6 | Primer selection | Provide primer selection and DNA amplification methods as well as variable region sequenced (if applicable). | | yes | yes | no | yes | yes | yes | NA | no | yes | no | yes | yes | yes | NA | NA | yes |
| 4,7 | Positive Controls | Describe any positive controls (mock communities) if used. | If used, should be deposited under guidance provided in the 8.X items. | not used | not used | not used | not used | not used | not used | not used | not used | not used | not used | not used | not used | not used | not used | not used | not used |
| 4,8 | Negative Controls | Describe any negative controls if used. | If used, should be deposited under guidance provided in the 8.X items. | not used | not used | not used | not used | not used | not used | not used | not used | not used | not used | not used | not used | not used | not used | not used | not used |
| 4,9 | Contaminant mitigation and identification | Provide any laboratory or computational methods used to control for or identify microbiome contamination from the environment, reagents, or laboratory. | Includes filtering of reagents and other steps to minimize contamination. It is relevant to state whether the specimens of interest have low microbial load, which makes contamination especially relevant. | not used | not used | not used | not used | not used | not used | not used | not used | not used | not used | not used | not used | not used | yes | not used | not used |
| 4,10 | Replication | Describe any biological or technical replicates included in the sequencing, including which steps were replicated between them. | Replication may be biological (redundant biological specimens) or technical (aliquots taken at different stages of analysis) and used in extraction, sequencing, preprocessing, and/or data analysis. | not used | not used | not used | not used | not used | not used | not used | not used | not used | not used | not used | not used | not used | not used | not used | not used |
| 4,11 | Sequencing strategy | Major divisions of strategy, such as shotgun or amplicon sequencing. | For amplicon sequencing (for example, 16S variable region), state the region selected. State the model of sequencer used. | yes | yes | yes | yes | yes | yes | yes | yes | yes | yes | yes | yes | yes | yes | yes | yes |
| 4,12 | Sequencing methods | State whether experimental quantification was used (QMP/cell count based, spike-in based) or whether relative abundance methods were applied. | These include read length, sequencing depth per sample (average and minimum), whether reads are paired, and other parameters. | no | no | yes | yes | yes | no | no | no | yes | yes | yes | no | no | no | yes | yes |
| 4,13 | Batch effects | Detail any blocking or randomization used in study design to avoid confounding of batches with exposures or outcomes. Discuss any likely sources of batch effects, if known. | Sources of batch effects include sample collection, storage, library preparation, and sequencing and are commonly unavoidable in all but the smallest of studies. | no | no | no | no | no | no | no | no | no | no | no | no | no | no | no | no |
