## Supplemental Table 5 for "A systematic review exploring the association between the human gut microbiota and brain connectivity in health and disease"

**The quality assessment of the MRI measurement per study, using a checklist adapted from the guidelines published by Poldrack et al. (2008)***

| | Curtis (2019) | Cai (2021) / Zhu (2022) / Zhang (2022) | Hall (2021) | Kohn (2021) | Tillisch (2017) | Gao (2019) | Kelsey (2021) | Wang (2019) / Zheng (2020) | Dong (2020) | Dong (2022) | Strandwitz (2019) | Ahluwalia (2016) | Labus (2019) | Li (2022) | Hong (2021) | Jacobs (2021) |
|---|---|---|---|---|---|---|---|---|---|---|---|---|---|---|---|---|
| **MRI system** | yes | yes | yes | yes | yes | yes | yes | yes | yes | yes | yes | yes | yes | yes | yes | yes |
| **MRI acquisition** (volumes, pulse sequence type, FOV, matrix size, slice thickness, acquisition orientation, whole brain, order of acquisition, TE/TR/flip angle)** | no | yes | yes | yes | yes | no | yes | yes | yes | yes | no | yes | yes | yes | yes | yes |
| **Preprocessing and registration** (software with version number, order of preprocessing, quality control measures, motion correction co-registration, atlas information, coordinate space, localization of anatomical locations, smoothing) | yes | yes | yes | yes | yes | yes | yes | yes | yes | yes | yes | no | yes | yes | yes | yes |
| **ROI/network selection** (how defined, how was signal extracted) | yes | yes | yes | yes | yes | yes | yes | yes | yes | yes | yes | yes | yes | yes | yes | yes |
| **Connectivity analysis** (seed-based/ICA? Toolbox used? Transformations? Voxel-wise correction?) | yes | yes | yes | yes | yes | yes | yes | yes | yes | yes | yes | yes | yes | yes | yes | yes |

*Poldrack, R. A., Fletcher, P. C., Henson, R. N., Worsley, K. J., Brett, M., & Nichols, T. E. (2008). Guidelines for reporting an fMRI study. Neuroimage, 40(2), 409-414.
**rs-fNIRS acquisition in the case of Kelsey (2021)
