## Supplemental Table 6 for "A systematic review exploring the association between the human gut microbiota and brain connectivity in health and disease"

**The quality assessment, quality rating and explanatory notes for each study, based on the criteria presented in table S3-5**

| Author (year) | Q1 | Q2 | Q3 | Q4 | Q5 | Q6 | Q7 | Q8 | Q9 | Q10 | Q11 | Q12 | Q13 | Q14 | Q15 | Q16 | Quality rating | | Notes |
|---|---|---|---|---|---|---|---|---|---|---|---|---|---|---|---|---|---|---|---|
| **Curtis (2019)** | yes | no | yes | no | NA | no | no | no | yes | no | no | no | NA | NA | no | no | 27% | Poor | 2. Unclear where and in what time period participants were recruited<br>4. Inclusion/exclusion criteria not consistent across groups and no inclusion criteria provided for the control participants<br>6. No sample size justification or reporting of effect size estimates<br>10. Lack of clarity regarding laboratory site, shipping, storage and sequencing methods<br>11. The gut microbiota was measured only once<br>12. Lack of detail regarding MRI acquisition<br>15. Analyses were only corrected for smoking status<br>16. Unclear which statistical tests were performed, and direction of effects not consistently reported |
| **Cai (2021) / Zhu (2022) / Zhang (2022)** | yes | no | yes | yes | NA | no | no | no | yes | no | no | yes | NA | NA | yes | yes | 64% | Fair | 2. Unclear where and in what time period participants were recruited<br>6. No sample size justification or reporting of effect size estimates<br>10. Lack of clarity regarding sequencing methods<br>11. The gut microbiota was measured only once |
| **Hall (2021)** | yes | no | yes | yes | NA | yes | no | no | yes | no | no | yes | NA | NA | yes | no | 64% | Fair | 2. No time-period for recruitment reported<br>10. The used primer is not reported<br>11. The gut microbiota was measured only once<br>16. No correction for multiple testing |
| **Kohn (2021)** | yes | no | yes | yes | NA | no | no | no | yes | yes | no | yes | NA | NA | no | yes | 64% | Fair | 2. Unclear where and in what time period participants were recruited<br>6. No sample size justification or reporting of effect size estimates<br>11. The gut microbiota was measured only once<br>15. The analyses were not corrected for age and sex, but the sample included only females in a narrow age-range |
| **Tillisch (2017)** | yes | no | yes | yes | NA | no | no | no | yes | yes | no | yes | NA | NA | yes | no | 64% | Fair | 2. Unclear where participants were recruited<br>6. No sample size justification or reporting of effect size estimates<br>11. The gut microbiota was measured only once<br>16. Methodological decisions not well described in the context of the sample size |
| **Gao (2019)** | yes | no | no | yes | NA | no | no | no | yes | no | no | no | NA | NA | yes | yes | 45% | Poor | 2. No time-period for recruitment reported<br>3. The rs-fMRI scan was not successful for over 50% of the eligible infants<br>6. No sample size justification or reporting of effect size estimates<br>10. Lack of clarity regarding sequencing methods<br>11. The gut microbiota was measured only once<br>12. Lack of detail regarding MRI acquisition |
| **Kelsey (2021)** | yes | no | yes | yes | NA | yes | no | no | yes | yes | no | yes | NA | NA | yes | yes | 82% | Good | 2. No time-period for recruitment reported<br>11. The gut microbiota was measured only once |

| Author (year) | Q1 | Q2 | Q3 | Q4 | Q5 | Q6 | Q7 | Q8 | Q9 | Q10 | Q11 | Q12 | Q13 | Q14 | Q15 | Q16 | Quality rating | | Notes |
|---|---|---|---|---|---|---|---|---|---|---|---|---|---|---|---|---|---|---|---|
| **Wang (2019) / Zheng (2020)** | yes | no | yes | no | NR | no | no | no | yes | no | no | yes | no | NA | yes | no | 42% | Poor | 2. No time-period for recruitment reported<br>4. Unclear whether the inclusion/exclusion criteria are consistently applied across cases and controls. There are inconsistencies in the description of the inclusion/exclusion criteria across the two reports (based on the same sample)<br>6. No sample size justification or reporting of effect size estimates<br>10. Lack of clarity regarding laboratory site, shipping and sequencing methods<br>11. The gut microbiota was measured only once<br>13. The analyses were not performed blinded to the case/control status of the participants<br>16. Unclear which tests were corrected for multiple testing |
| **Dong (2022)** | yes | no | yes | yes | yes | no | no | no | yes | no | no | yes | no | NA | no | yes | 58% | Fair | 2. Unclear where and in what time period participants were recruited6. No sample size justification or reporting of effect size estimates10. Lack of clarity regarding laboratory site, sample collection and shipping11. The gut microbiota was measures only once13. The analyses were not performed blinded to the case/control status of the participants15. The analyses were not corrected for age and sex |
| **Dong (2020)** | yes | no | yes | yes | NA | no | yes | yes | yes | no | no | yes | NA | yes | no | no | 57% | Fair | 2. No time-period for recruitment reported<br>6. No sample size justification or reporting of effect size estimates<br>10. Lack of clarity regarding laboratory site, sample collection, shipping, storage and primers used<br>11. The gut microbiota was measured only once before and after the intervention<br>15. The analyses were not corrected for age and sex<br>16. Lack of clarity regarding methodological decisions. E.g., it is not explained why data from different timepoint was pooled |
| **Strandwitz (2019)** | yes | no | yes | yes | NA | no | no | no | yes | yes | no | no | NA | NA | yes | yes | 64% | Fair | 2. No time-period for recruitment reported<br>6. No sample size justification or reporting of effect size estimates<br>11. The gut microbiota was measured only once<br>12. Lack of detail regarding MRI acquisition |
| **Ahluwalia (2016)** | yes | no | yes | yes | NA | no | no | no | yes | no | no | no | NA | NA | no | no | 36% | Poor | 2. No time-period for recruitment reported<br>6. No sample size justification or reporting of effect size estimates<br>10. Lack of clarity regarding laboratory site, sample collection, shipping, storage and sequencing method<br>11. The gut microbiota was measured only once<br>12. MRI pre-processing steps not reported<br>15. The analyses were not corrected for age and sex<br>16. Lack of clarity regarding statistical approach |
| **Labus (2019)** | yes | yes | yes | yes | NR | no | no | no | yes | no | no | yes | no | NA | yes | no | 58% | Fair | 6. No sample size justification or reporting of effect size estimates<br>10. Lack of clarity regarding sample storage and sequencing method<br>11. The gut microbiota was measured only once<br>13. The analyses were not performed blinded to the case/control status of the participants<br>16. No correction for multiple testing |

| Author (year) | Q1 | Q2 | Q3 | Q4 | Q5 | Q6 | Q7 | Q8 | Q9 | Q10 | Q11 | Q12 | Q13 | Q14 | Q15 | Q16 | Quality rating | | Notes |
|---|---|---|---|---|---|---|---|---|---|---|---|---|---|---|---|---|---|---|---|
| **Li (2022)** | yes | no | yes | yes | NR | yes | no | no | yes | no | no | yes | no | NA | no | no | 50% | Fair | 2. No time-period for recruitment reported<br>10. Lack of clarity regarding pre-processing steps<br>15. The analyses were not corrected for age and sex<br>16. Lack of clarity regarding which statistical tests were used and which variables were used for functional connectivity |
| **Hong (2021)** | yes | yes | yes | yes | NA | no | yes | yes | yes | yes | no | yes | NA | no | yes | yes | 79% | Good | 6. No sample size justification or reporting of effect size estimates<br>11. The gut microbiota was measured only once before and after the intervention<br>14. Loss to follow-up was more than 20% |
| **Jacobs (2021)** | yes | yes | yes | yes | NA | no | yes | yes | yes | yes | no | yes | no | yes | yes | yes | 80% | Good | 6. No sample size justification or reporting of effect size estimates<br>11. The gut microbiota was measured only once before and after the intervention<br>13. The analyses were not performed blinded to the response status of the participants |

NA: not applicable; NR: not reported

| | |
|---|---|
| **Poor (<50%)** | **4** |
| **Fair (50-74%)** | **9** |
| **Good (≥75%)** | **3** |
